# Sporadic, late-onset, and multistage diseases

**DOI:** 10.1101/2021.12.15.21267843

**Authors:** Anthony J. Webster, Robert Clarke

## Abstract

Somatic mutations can cause cancer and have recently been linked with a range of non-malignant diseases. Multistage models can characterise how mutations lead to cancer, and may also be applicable to these other diseases. Here we found the incidence of over 60% of common diseases in UK Biobank were consistent with a multistage model with an ordered sequence of stages, as approximated by a Weibull distribution, with the log of incidence linearly related to the log of age and the slope often interpreted as the number of stages. A model where the stages can occur in any order was also explored, as was stratification by smoking and diabetes status. Most importantly, we find that many diseases are low risk when young but then become inevitable in old age, but many other diseases do not, being more sporadic with a modest and modifiable risk that slowly increases with age.

---

Many common diseases are thought to have a multistage aetiology [1–12], possibly involving one or more genetic mutations [5, 13–18]. One of the most important shifts in our understanding in recent decades, is the discovery that somatic mutations are prevalent in the majority of tissues in our body [14, 19–22]. The best known examples include clonal expansions that co-exist with healthy cells in our blood [14, 15], skin [19], and esophagus [20]. This raises the possibility that somatic or epigenetic mutations could contribute to the initiation of other non-cancerous diseases [13–18, 23]. This includes generic processes such as inflamaging [24, 25], and entire classes of diseases, including autoimmune and cardiovascular diseases [13–18, 23]. The involvement of mutations in autoimmune diseases was suggested 50 years ago [23], and can now be explored in detail. If mutations are involved in triggering disease, then it is likely that they might constitute one or more rate-limiting steps for disease onset, in a similar way to cancer incidence.

We assessed the potential frequency of multistage processes in the most common diseases in UK Biobank, as could arise from a sequence of rate-limiting processes such as somatic mutations. A total of 800 diseases were studied using two simple parametric models of multistage disease processes, both of which accurately described the incidence rates for approximately 500 of the diseases considered, In addition to potential biological insights, the parameterised models provide important new insights into disease incidence, that were not apparent from conventional studies using relative risks. We identified late-onset diseases that appear to become inevitable at the extreme ends of observed human lifespan, and those that are more sporadic, often occurring at younger ages, but only weakly influenced by age and potentially avoidable. The incidence of these “sporadic” diseases appears to be particularly sensitive to lifestyle interventions and modifiable risk factors, suggesting that their incidence may be substantially reduced.

One striking observation was that “all diseases are rare”, in the sense that without germline genetic abnormalities the probability of surviving a particular disease *S*(*t*) for a typical human lifetime, is never much less than 1. Living to old age is unlikely because of the large number of potentially fatal diseases, the risk of any single disease is comparatively low. As a result, it is often reasonable to approximate the probability density for disease incidence *f* (*t*) by its hazard function [26], with *f* (*t*) ≃ *h*(*t*), and the survival function *S*(*t*) ≃ 1. This simplifies the construction and interpretation of models for the incidence of disease.

## Multistage models

The multistage (or “multistep”) model was inspired in the 1950s by a biological model in which cancer involves several genetic mutations before symptoms are observed [1–3, 27]. The models describe any process that can arise through one or more independent pathways, with one or more sequential or non-sequential steps [26] (figure 1). Whereas proportional hazards models are ideal for studying associations with risk factors, multistage models are ideal for characterising incidence rates. They provide a concept of relative aging rate, that for a specific disease, captures the relative ages of exposed compared to unexposed individuals. Other advantages include an interpretation in terms of rate-limiting processes, that if biological in origin, might be targeted to slow or prevent disease. They also allow incidence rates to be predicted and extrapolated beyond the observed data, allowing unique insights about disease incidence that cannot be captured by histograms or proportional hazards studies ^1^.

**Figure 1:**
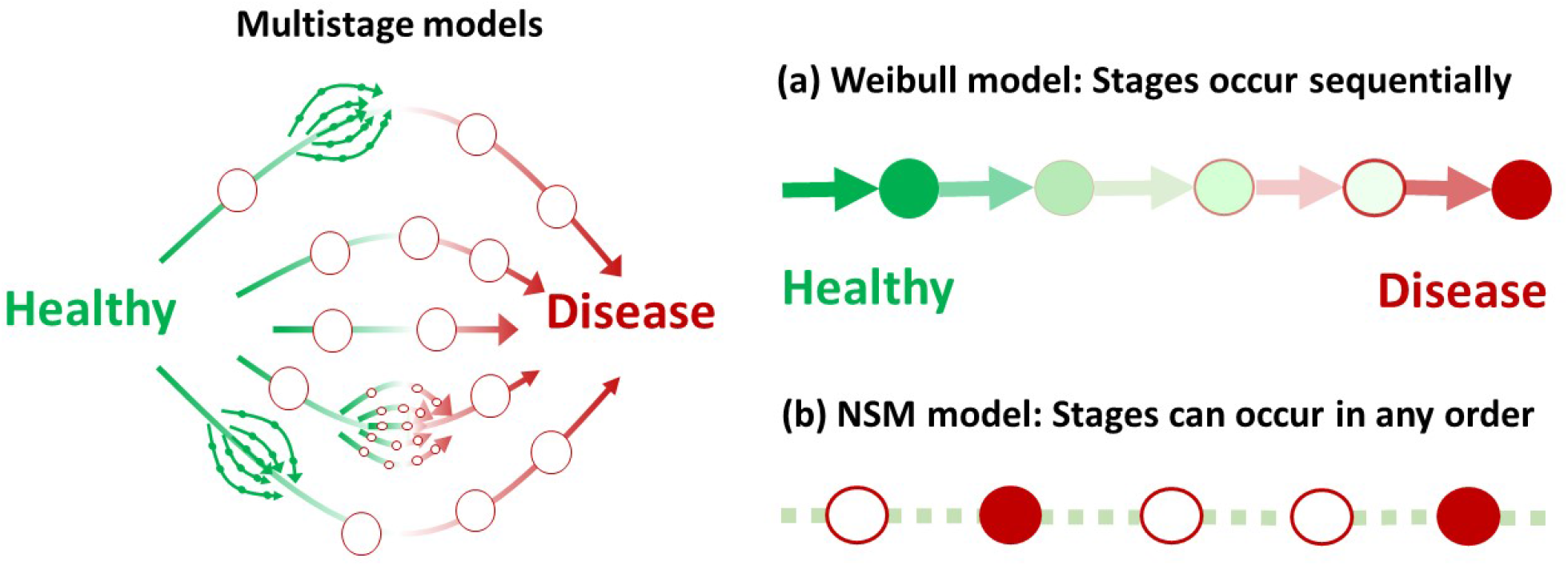
A multistage model can describe any process that arises through one or more independent pathways, with one or more sequential or non-sequential steps. (a) The sequential Armitage-Doll (Weibull) model requires that stages occur sequentially. (b) The non-sequential model (NSM), allows the stages to occur in any order.

Two models are considered here (figure 1a and 1b). The first approximates a sequential multistage model of disease (figure 1a), with a Weibull model. This approximation is good when diseases are rare over a typical human lifetime, as is the case here. The Weibull model has the benefit of being a proportional hazards form, allowing us to estimate associations with potential risk factors *X* in the conventional way, with a hazard function *h*(*t*) = *e*^*βX*^ *h*_0_(*t*). giving a survival function,

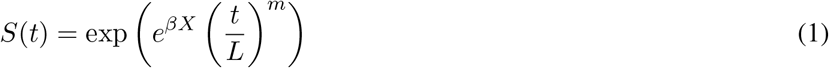

The exponent of *S*(*t*) is referred to as the cumulative hazard function *H*(*t*) = ∫^*t*^ *h*(*s*)*ds*, and can be rearranged as:

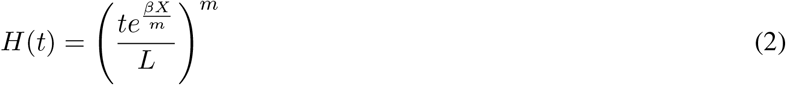

This gives an effective age 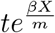, that is determined by risk factors *X*, with the factor 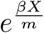 giving a relative aging rate compared with the baseline values where *βX* = 0 and 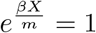. It also emphasises that for any given disease, the influence of risk factors on your aging rate through *βX*, will be suppressed by a factor of 1*/m*, that will be smaller for diseases with many steps *m*.

The second model considers multistage processes that can occur in any order (figure 1b), and takes all events to happen with approximately constant rates. This is a reasonable approximation, because within a non-sequential model all the rate-limiting steps must have similarly slow rates, or they would not be rate-limiting and observable from the incidence data. This is discussed further in the Supplementary Material. We refer to this non-sequential model as the “NSM” model, and its survival function is (see Supplementary Material),

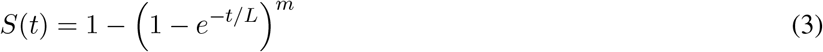

where *m* is the effective number of steps, *t* is age, and *L* is a time scale for the disease processes. This model has a very different biological interpretation than the Weibull model, typically with a much larger number of steps.

The Weibull and NSM models are derived in the Supplementary Material, along with several relevant observations that have not been published elsewhere. One (important) counter-intuitive observation, is that even when a disease proceeds sequentially through a series of *m* steps, if the rate of one or more steps increase sufficiently, then the observed number of steps will decrease, despite the associated biological processes continuing to occur. Although not necessarily obvious, the mathematical result is very clear. Another observation is that it will often be reasonable to study multistage disease processes with a proportional hazards model. If the hazard of every stage can individually be approximated by a proportional hazards model, then for the “rare” diseases that we study here, a multistage disease can also be described by a proportional hazards model. However, the estimated associations are *β* = Σ_*i*_ *β*_*i*_, a sum over the associations at each step, and each *β*_*i*_ could be quite different. This is particularly important for sequential disease processes, because it emphasises that the same risk factor could have a different influence at different stages of the disease, that in practice could produce different associations at different ages. A final observation is that because individual diseases are rare over a human lifetime and the incidence of diseases that are studied are similar, ranging between a few hundred to a few thousand cases, the hazard of the observed diseases must be a similar order of magnitude. This requires (*t/L*)^*m*^ to be similar for all diseases where *t* ∼ 60, and leads to *m* and *L* being strongly correlated, with *m* log(*L*) being similar for the diseases studied here.

## Results

### Goodness of fit consistency with multistage models of disease

The Weibull and NSM models both accurately fitted the incidence of most of the 800 diseases considered, with 485 with Weibull, 466 with NSM, and 450 diseases included in both sets (see Supplementary Material for details and example plots). For most diseases, despite the similarly good fit to the data by both the Weibull and NSM models, they require different numbers of steps in a multi-stage model of disease. This demonstrates that a good fit to the data with a multi-stage model, cannot on its own, be taken as evidence for an underlying multi-stage biological process. The standard error of the effective number of steps was greater than 0.35 for 365 (417) of the fitted diseases for the Weibull (NSM) models respectively, making it unlikely to see clear indications of integer-valued *m* for either model. A plot for the fitted values of the effective number of steps *m*, ordered by *m*, showed no visual evidence for steps at integer-values (Supplementary Material, figure 6). Neither was there any evidence for a systematic reduction in *m* for smokers, as you would expect if smoking reduced the number of steps needed to trigger disease (Supplementary Material, figure 7).

### Sporadic versus late-onset disease

Parametric models for disease incidence allow comparisons of different patterns of disease incidence (first occurrence of disease in each ICD-10 chapter). Only diseases whose incidence data were sufficiently well fitted by a multistage model were considered (as discussed above). Figure 2 shows the cumulative probability distribution function for disease by age 50, denoted *F* (50), versus the relative increase in disease risk by age 100, (*F* (100) − *F* (50))*/F* (50). The probabilities were estimated with a Weibull model, and the NSM model gave almost identical results (see Supplementary Material). Diseases with a late age of onset are near the top of the figure, where disease risk increases rapidly at older ages. These contrast with diseases near the bottom of the figure, whose risk increases comparatively slowly. Diseases with the lowest risk at age 50 are towards the figure’s left, and those with highest risk are towards the figure’s right. Diseases at the bottom right, have a comparatively high probability of occurrence by age 50 years (*F* (50) was larger than for most diseases), and the increased probability of disease at age 100 compared to age 50 is comparatively low (*F* (50)*/F* (100) is higher than for most diseases)^2^. Therefore we distinguish between the most “sporadic” and “late-onset” diseases by the product *F* (50) × *F* (50)*/F* (100), that is largest (smallest), for the most (least) sporadic diseases. The diseases were classified into tertiles in figure 2, with most sporadic (orange), mid-range (green), and late-onset (red). The classification enabled a qualitative comparison between the late-onset and sporadic diseases.

**Figure 2:**
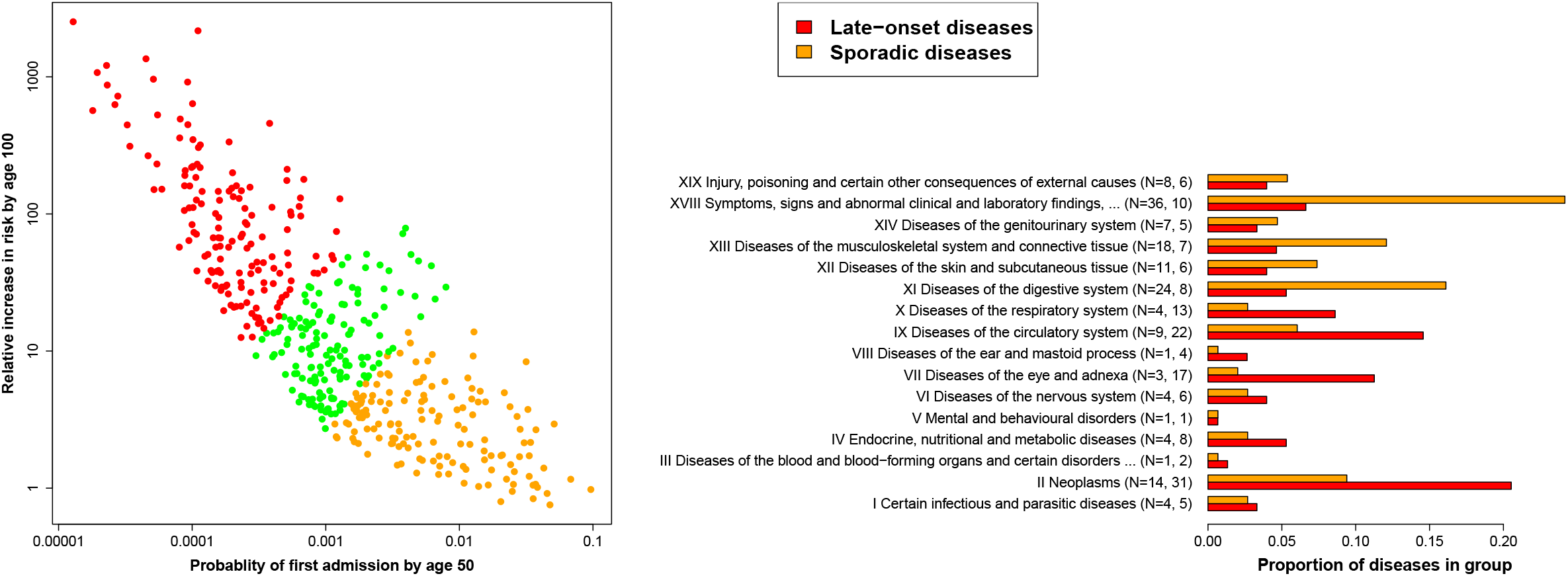
Left: For each disease, the probability of first hospital admission at 50 is plotted versus the relative increase in probability of disease by age 100. Disease incidence was classified in tertiles to indicate how sporadic or late-onset it was (see main text). Results for the Weibull model are shown, the NSM model gave similar results (Supplementary Material). Right: The composition of sporadic and late-onset diseases in terms of ICD-10 chapters, with N the number of diseases.

Figure 2 also shows the “sporadic” and “late-onset” diseases in terms of ICD-10 chapters. The late-onset diseases are composed primarily of: II Neoplasms, VII Diseases of the eye and adnexa, XI Diseases of the circulatory system, and X Diseases of the respiratory system. In contrast the more sporadic diseases are mainly composed of: XI Diseases of the digestive system, XIII Diseases of the musculoskeletal system and connective tissue, XVIII Symptoms, signs and abnormal clinical and laboratory findings, not elsewhere classified. Similar results were found when men and women were considered separately (see Supplemental Material, Section D).

### Relative aging rates, and modifiability of risk

The Weibull model has the form of a proportional hazards model, which makes adjustment for variables such as smoking status, easy to interpret. The Weibull model also allows the definition of a relative aging rate *e*^*βX/m*^ (see Eq. 2), that when multiplied by your age, gives your effective age for each disease in terms of your risk factors *X*, using the estimated coefficients *β* and exponent *m* for each disease. This provides an alternative measure to relative risk. Whereas relative risk measures your risk at a given time relative to the baseline, the relative aging rate allows your effective age to be determine in terms of your risk factors.

Figure 3 considers sporadic and late-onset diseases, and shows box plots for relative risks and relative aging rates. Comparing relative risks, the influence of diabetes and smoking were more important for strongly age-related diseases, but the results are otherwise similar. In contrast, the relative aging rates estimated for sporadic diseases were substantially larger than for diseases with a late-onset, with the relative aging rates associated with minimum education, maximum BMI, and diabetes, all being of order 1.1, and diseases with a late-onset typically less than half that. A relative aging rate of 1.1 would indicate that someone aged 50 would be at equivalent risk to someone aged 55 years with baseline risk factors. For a diabetic, in the maximum BMI tertile, and minimum education group, the relative aging rate could easily be 1.3, indicating that someone aged 50 years would be at equivalent risk to someone aged 65 without these risk factors, or someone aged 70 would be at equivalent risk to someone aged 91. In terms of disease-free years [29], if someone with baseline risk factors aged 40 was expected to have 25 disease-free years, then someone with a relative aging rate of 1.1 would on average reach (40+25)/1.1≃59 years before their first disease (19 disease-free years from age 40).

**Figure 3:**
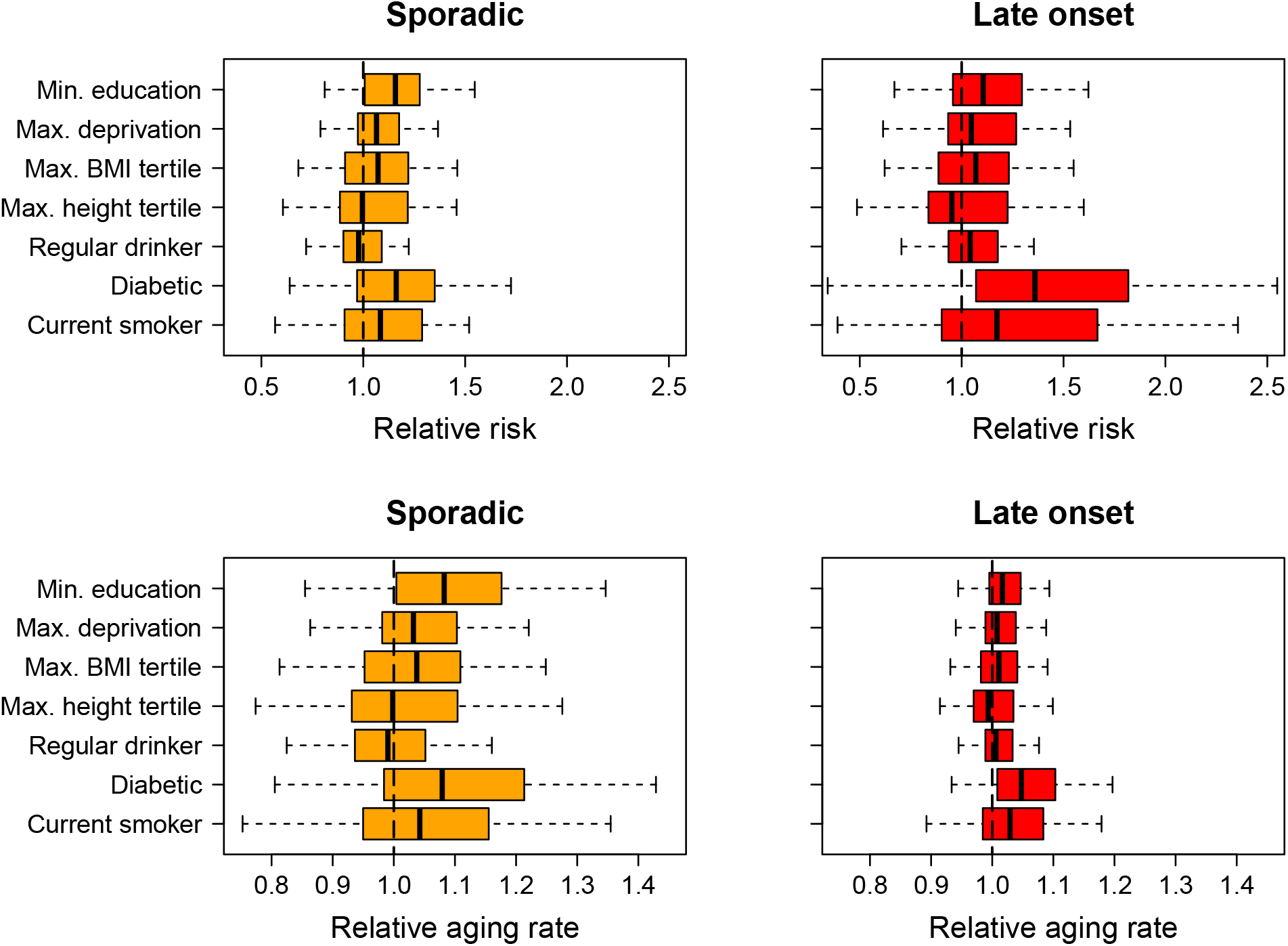
Relative risks (top), and relative aging rates (bottom), for potential risk factors associated with sporadic diseases (left), and late-onset diseases (right). Box plots show the median, the interquartile range, and whiskers at 1 × the interquartile range. Relative risks for diabetes and smoking tended to be larger for late-onset disease. Relative aging rates tended to be larger for sporadic diseases, suggesting that your effective age at risk is more modifiable for sporadic than late-onset diseases.

### Stratification by smoking status and diabetes

Qualitative information about the influence of an exposure or risk factor on disease can be obtained by stratifying data by e.g. smoking status, and understanding how to plot and interpret the data. For the cumulative hazard function of a Weibull distribution *H*(age), when adjusted for the survival function being less than 1 at a participant’s entry into the study, a plot of log(*H*(age)) versus log(age)) will appear as a straight line (figure 4 and Eq. 2). If the Weibull distribution represented a multi-stage model of disease, and the rates of one or more stages in the disease process were increased, then the plot would be displaced vertically upwards. If the rate of one or more stages increased sufficiently that they were no longer a rate-limiting step (Section B of Supplementary material), then the plot would be displaced vertically upwards and reduced in slope. If the rates of each stage were being decreased, then the opposite would happen, with a vertical displacement of the plot downwards and possibly associated with an increase in slope if the rates of one or more stages were decreased sufficiently. Therefore if we stratify by strong risk factors such as smoking or diabetes status, then we can explore how these risk factors qualitatively modify disease risk under the hypothesis of an underlying multistage model (figure 4, and figures 3-s5 in the Supplementary Material).

**Figure 4:**
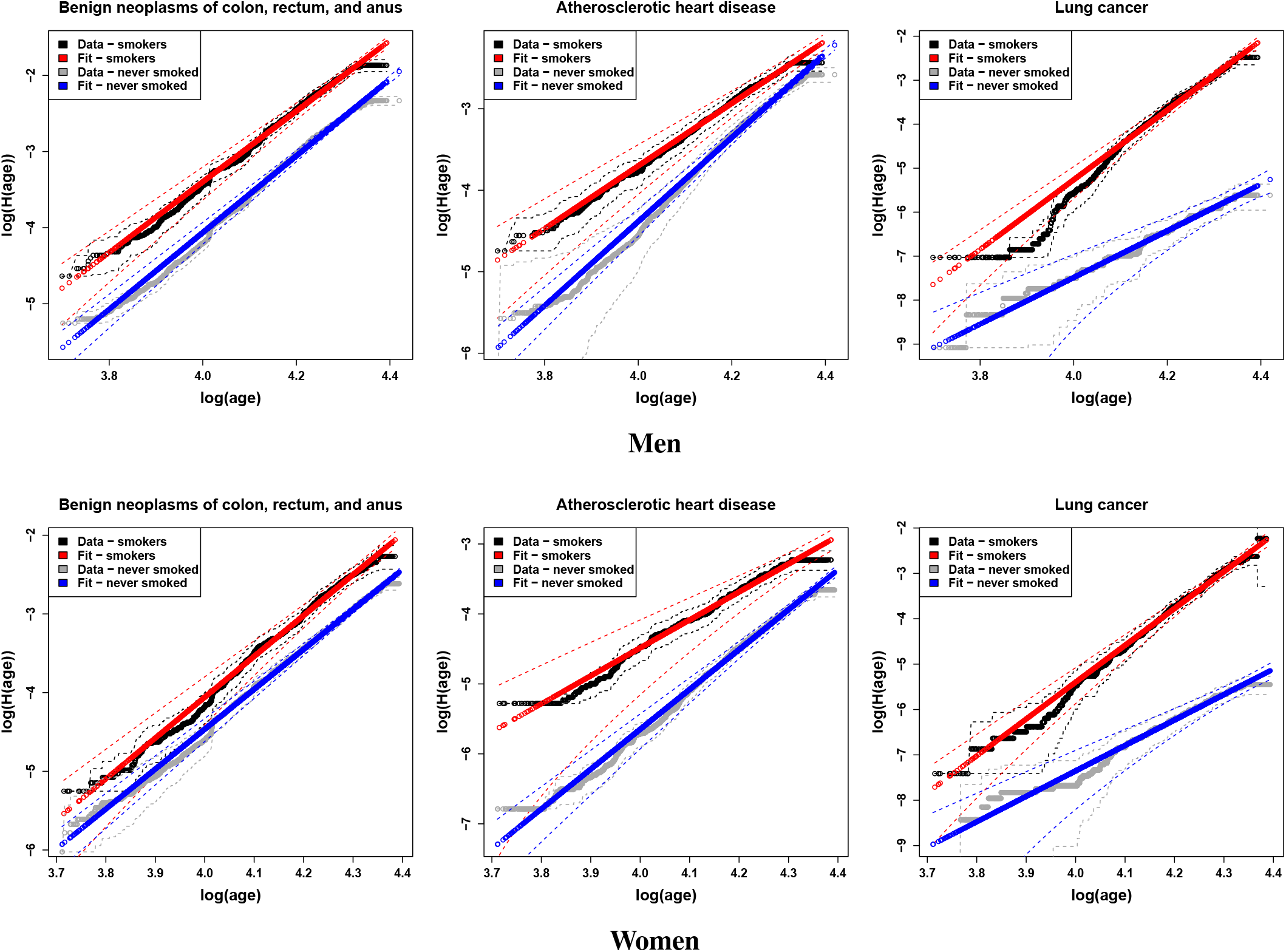
The incidence of three common smoking-related diseases in men (top) and women (bottom), stratified by smoking status, with Kaplan-Meier estimates adjusted using Eq. 5. Data for smokers differ by vertical displacements and changes in slope, that can be interpreted as changes in the rates of processes, and numbers of steps in disease (see main text).

The influence of smoking was strongly disease-dependent. Figure 4 illustrates the differences. For benign bowel cancers (D12), the vertical displacement of plots between smokers and non-smokers is consistent with smoking increasing the rate of one or more processes that are leading to the disease, and the stratified data appear as parallel lines. For atherosclerotic heart disease (I25.1), the increased rate and reduced slope for smokers is consistent with smoking leading to an effective reduction in the number of steps needed to trigger disease, and the stratified data appear as lines that converge with age. However, for lung cancer (C34), the disease that is most strongly influenced by smoking, both the rate and slope of the curve increase, with the stratified datasets diverging with age. Reasons for this unexpected increase in slope (effective number of steps), that are specific to lung cancer, are discussed later.

Disease risks associated with diabetes could be driven by shared genetic or other risk factors, or mediated by diabetes. As an example, three common cardiovascular diseases affecting men and women were considered (Supplementary Material figure 3). Atherosclerotic heart disease and atrial fibrillation are consistent with a multistep disease process in which increases the rates of disease processes, possibly sufficiently to reduce the number of rate-limiting steps needed to trigger disease. For pulmonary embolism, the rates were similar in diabetic and non-diabetic women, but there was evidence for a reduction in risk of pulmonary embolisms (I26) in the strata of diabetic men.

## Discussion

### Prevalence of multistage disease processes

Approximately two thirds of the diseases considered were consistent with a multi-stage model of disease, in which diseases require several distinct events, or a sequence of stages before the disease is observed. However, several authors have remarked [26] that the Weibull model in particular, would be expected to fit a wide range of models. Both the Weibull and NSM models provided good fits to the data, despite their mathematical differences and distinct biological interpretations. Therefore, based on goodness of fit to incidence data alone, it is unlikely that all the diseases have an underlying multistage aetiology.

### Stratification by smoking status or diabetes

Smoking is known to increase the risk of many diseases, especially cancer and cardiovascular disease. If a disease develops through a multistage process, then exposure to smoking might be expected to change the rates of one or more disease processes. As demonstrated mathematically in Section B of the Supplementary Material, if the rate of any given stage increased sufficiently, then exposure to smoking can reduce the effective number of rate-limiting steps *m* observed in the incidence data. Therefore we might (wrongly) expect that if smoking was increasing the rate of one or more disease processes, then *m* should either decrease or remain unchanged. This is certainly what is seen for benign bowel cancers (D12) and bladder cancer (C67), in figure 4, with a vertical upward displacement of the curves indicating an increased rate of processes, and for heart disease (I25.1), that shows an increased rate and a decreased slope (decreased number of steps *m*). In these cases, the interpretation in terms of multistage processes is compelling, and may warrant further investigation.

In contrast, the influence of smoking on lung cancer incidence was initially surprising. Both the rate, and the slope (effective number of steps *m*), increase (figure 4). It is possible that the influence of cigarette smoke was so strong that it is entirely changing the landscape of rate-limiting steps that are needed to trigger disease. For example, if the rates of the *previously* rate-limiting steps were increased sufficiently, then there would be an entirely new set of rate-limiting processes that would determine the observed incidence data. This could produce an increased rate and an increase in *m*. Importantly, lung cancer has several different types, that within ICD-10, are grouped into a single composite endpoint denoted by code C34 that was studied here. It is known that the proportions of adenocarcinoma are much higher in non-smokers than smokers, with smoking more rapidly increasing the risk of other types of lung cancer. It seems likely that different subtypes of diseases are influencing, and possibly dominating, the incidence patterns of lung cancer in smokers. This would provide one biological manifestation of a scenario that could substantially change the landscape of rate limiting processes, and the resulting incidence of disease.

If smoking is increasing the risks of different cancers to those in non-smokers, then either different processes such as different patterns of mutation in cells are occurring, or cancer is being triggered in different cell types. A recent study of somatic mutations in the bronchial epithelium found that cells with high rates of mutations in previous smokers, are replenished over time with near-normal cells, and hypothesised that there is an ongoing replacement of progenitor cells from a pool of quiescent stem cells [30]. If cancer initiation were solely in progenitor cells, then it would involve a competing-risk process between the cell acquiring sufficient mutations to form a cancer, and the cell’s death. Cancer risk in smokers would then be expected to saturate with age at a level where cells are being regularly replaced, and the statistical burden of mutations among cells was similar, unless there is a stage where cell death is avoided due to e.g. a prior somatic mutation. Cells could then remain in a precancerous state, until one or more mutations caused them to develop into a cancer at a potentially much later age. The role of smoking would be to increase the rate of these set of disease processes sufficiently that it becomes the dominant observed type of lung cancer. These remarks are consistent with the suggestion that smoking may modify the rate constants for both an early and a late-stage in a multistage model of lung cancer, but not all of them [3].

The differences in incidence rates between diabetics and non-diabetics were generally consistent with the hypothesis that diabetes increases the rates of disease processes (figure 3 in the Supplementary Material), and this can be sufficient to reduce the effective number of steps prior to disease. An alternate explanation is that one or more genetic risk factors for diabetes are acting to reduce the effective number of steps needed to trigger disease. An interesting anomaly is pulmonary embolism in men, for which the incidence rate appears to be substantially reduced in diabetics. In contrast, for women there were no differences between the diabetic and non-diabetic survival curves. There is no adjustment for other factors in the plots, just stratification by diabetes status at entry to the study. Nonetheless, the differences warrant further investigation.

### Comparison with conventional methods

Differences in incidence data can be identified by comparing conventional Kaplan-Meier survival curves, but survival curves would not appear as straight lines, and the differences would not have any particular interpretation. Equivalent plots to figure 4 require adjustment for the survival function being less than 1 at a participant’s entry into the study, for example, using a Weibull model for the adjustment. Differences between stratified data can subsequently be interpreted in terms of changes to the rates of processes or the number of steps, in an assumed multi-stage model of disease. Irrespective of whether the data reflect an underlying mechanistic process with distinct biological steps, it can be helpful to identify when the data can be modelled with several rate-limiting steps, providing a conceptual (hypothetical) model for disease onset that can be tested and explored.

Proportional hazards models are often used to estimate associations with risk factors. The proportional hazards assumption allows adjustments that correspond to vertical displacements of the curves in figure 4. Therefore, if adjusting for smoking with a proportional hazards model and the data considered here, figure 4 suggests that the model could accurately describe benign bowel cancers (D12), and approximate atherosclerotic heart disease (I25.1) or acute myocardial infarctions (I21), but it would be a poor approximation for lung cancer (C34). Whereas these differences are clear in figure 4, they may not be clear from a statistical test of the proportional hazards assumption.

### Can sporadic disease be avoided?

The risk of late-onset, strongly age-related diseases, increases sufficiently rapidly with age to make them almost inevitable at the extreme end of observed human lifespan. In contrast, even at the end of a typical human lifespan, there appears to be a reasonable statistical chance of having avoided the more sporadic diseases entirely. In addition, the risk of the more sporadic diseases, tends to be the most modifiable by established risk factors when measured in terms of relative aging rates. For example, someone with diabetes in the lowest education and maximum BMI tertiles, has an equivalent age at risk of the “sporadic” diseases that is about 30% higher than someone at baseline. This would be expected to increase rates of hospitalisation, irrespective of disease-disease interactions, or multimorbidity. From a statistical perspective, the results suggest that lifestyle interventions or appropriate medications might substantially reduce the incidence of a large proportion of these more sporadic diseases, possibly for an entire lifetime.

An important distinction between the diseases considered here, is whether they are curable. Whereas many of the diseases can be treated with surgery, drugs, or lifestyle interventions, diseases such as cancers have a much less certain longer-term prognosis. The sporadic diseases seem likely to have the biggest potentially avoidable impact on general health and health costs, but if you are unlucky enough to get an incurable disease then this is much more serious for the individual involved. This must be bourne in mind when deciding whether to focus extra effort on sporadic diseases to reduce hospital admissions.

### What can we learn from parametric and multistage models?

The main advantages of multistage models are their ability to quantify the age-dependence of disease incidence, and their provision of a simple conceptual model for the progression of disease. Because two different models with very different biological interpretations, both describe the data well, any biological interpretation of the results should be treated with caution. However, the models provide a helpful picture of disease onset, that appears to have been valuable within cancer research. They also encourage us to think more carefully about modelling disease. For example, Section B of the Supplementary Material observes that a sequential progression of disease through stages could have age-dependent differences in associations, if the associations differed between e.g. an earlier and a later stage. Early in cancer research, a stage-dependent influence of exposures was suggested for smoking and lung cancer risk [3], but such issues are rarely considered in epidemiology. The clearest advantage of the models as used in this study, is the parametric description of disease incidence, that provided new perspectives on the age-dependent causes of hospital treatment. Specifically, by being able to estimate disease risk at two very different ages, we were able to clearly demonstrate that statistically at least, some disease-risk increases rapidly with age, but others might be avoided entirely. The Weibull model’s relative aging rate also provides a more meaningful measure of risk modification than relative risk, with the former determining an effective age for each disease in terms of risk factors.

## Conclusions

Simple, biologically-inspired, parametric models of disease, were found to accurately approximate the incidence rates of approximately 60% of diseases in the UK Biobank dataset. Because the incidence data of most diseases were described equally well by two models with very different biological interpretations, we should be cautious about inferring that such diseases arise from a multistage process. The parametric models yielded two important new insights. The first was that the risk of some diseases increases rapidly with age, but others are more sporadic. Statistically at least, over a human lifetime, we might hope to avoid the more sporadic diseases entirely. Secondly, the Weibull model introduced the concept of relative aging rate, that provides an intuitive alternative to relative-risk for understanding how your likelihood of disease is modified by risk factors. The more sporadic diseases tended to have relative aging rates that were more modifiable by established risk factors than the diseases with a late-onset in life. Overall, the findings suggest that a substantial proportion of hospital admissions for the more sporadic diseases, might be avoided by adopting a healthier lifestyle.

## Methods

### Data sources

The UK Biobank dataset [31] was used, that involved over 500, 000 men and women aged between 49 and 69 years, who were recruited during 2006-2010. Primary diagnoses of diseases in hospital records were considered, that were recorded with an International Classification of Diseases version 10 code (ICD-10) [32, 33]. For data access, see “Data availability”.

### Diseases studied

The data set and disease definitions are described in detail elsewhere [31, 34]. Diseases were defined in terms of one or more 4-digit International Classification of Disease Codes (ICD-10), selected by two epidemiology-trained clinicians with specialities in pathology and general practice. The selected diseases were intended to be accurately diagnosed, distinct, and typically age-related causes of disease. Any coded diseases with an ambiguous underlying cause were excluded, as were diseases due to chance events or exposures such as an accident, or an infection while on holiday. To ensure that disease diagnoses had passed a threshold of severity, and were unlikely to result from undiagnosed or co-occurring disease, we focussed on diseases that were the primary diagnosis (and primary reason for hospital admission). As a compromise between avoiding confounding by prior disease, but retaining sufficient cases for a meaningful statistical study, we consider each individual’s first hospital admission for a disease in each ICD-10 chapter. As a result, there could be more than one case of disease for each individual, but the diseases will be from different ICD-10 chapters. Data were excluded if participants had a cancer other than non-melanoma skin cancer before their first assessment in the UK Biobank study (the start of the study period).

### Statistical analysis

The R software package was used for all analyses [35]. Age was used as the time variable, left-truncated at entry into the study, and right-censored at either the study end or following incidence of a cancer other than non-melanoma skin cancer. Maximum likelihood estimates for parameters were obtained by numerically maximising a left-truncated and right-censored log-likelihood for the data, using the “maxLik” package [36]. When there was adjustment for potential risk factors, initial estimates were calculated using a proportional hazards model and the “survival” package [37, 38]. To measure goodness of fit, Kaplan-Meier estimates to the data were compared with the parameterised models. Key to doing this correctly, is the observation that the Kaplan-Meier fit assumes *S* = 1 at the study’s start, whereas it may already be less than one if age is the time variable and participants join the study in middle-to old-age. The fitted survival functions were used to estimate *S*(*t*_1_) = exp(− *H*(*t*_1_)), where *H*(*t*) is the cumulative hazard function and *t*_1_ is the age at the first observed event, by firstly writing,

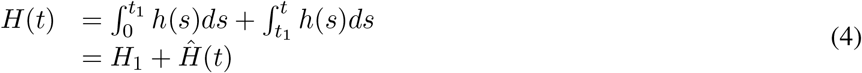

and noting that the Kaplan-Meier estimator approximates Ŝ = exp(− *Ĥ* (*t*)). Then with the parameterised model for *H*(*t*), and the Kaplan-Meier estimate for *Ĥ* (*t*), we can rearrange Eq. 4 and average over the data to estimate,

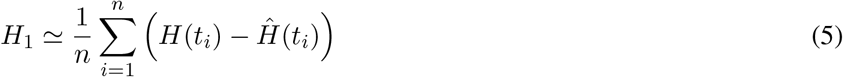

The Kaplan-Meier estimate can then be adjusted to account for *S ≠* 1 at the study’s start, with *S*(*t*) = exp(−*H*_1_) exp(*Ĥ* (*t*)). Without recognising the need for this adjustment, and doing so correctly, the incidence of most diseases in UK Biobank would appear very differently, and would wrongly suggest that multistage diseases are very rare. Note that there are no additional free parameters introduced by the estimate. The parameterised model was compared with the Kaplan-Meier estimator using standard *χ*-square tests, using the variance at each point from the Kaplan-Meier estimate, and (optionally) with the variance for the parameterised model fit estimated using the delta method [39].

All analyses were considered in men and women separately. For studies with adjustment, we considered common established risk factors of: diabetes (no, yes), smoking status (never, previous, or current), alcohol consumption (rarely – less than 3 times per month, sometimes - less than 3 times a week but more than 3 per month, regularly - 3 or more times each week), education (degree level, post-16 but below degree, to age 16 or unspecified). We considered tertiles of deprivation, and separately in men and women, tertiles of height and BMI. For women, we also adjusted for HRT use ever (yes, no), and one or more children (yes,no). Baseline was taken as: no diabetes, never smoker, rarely drink, brisk walking pace, degree-level education, minimum deprivation tertile, and women with no children or HRT use. Analyses were multiply adjusted. The data had less than 1% missing values, allowing a complete case analysis.

R packages used during the analysis, to manipulate data, and to create plots and tables include: maxLik[36], survFit[37, 38], grr[40], data.table[41], bit64[42], pracma[43], fmsb[44].

## Data Availability

UK Biobank data can be accessed by application through www.ukbiobank.ac.uk, and summary data produced during this study will become available from OSF after peer review is complete.

## Data availability

UK Biobank data can be accessed by application through www.ukbiobank.ac.uk, and summary data produced during this study will become available from OSF (https://osf.io), shortly. UK Biobank has approval by the Research Ethics Committee (REC) under approval number 16/NW/0274.

## Code availability

R code used to produce figures from summary data will become available with the summary data at OSF (https://osf.io), shortly. The full code for use with non-summary data will be returned with other results to UK Biobank (see www.ukbiobank.ac.uk).

## Acknowledgements

This research has been conducted using the UK Biobank resource under application number 42583. Anthony Webster is supported by an intermediate research fellowship from the Nuffield Department of Population Health (NDPH). Robert Clarke is supported through core funding for NDPH from the Medical Research Council (MRC) and the British Heart Foundation (BHF). The University of Oxford MRC Population Health Research Unit is funded through a strategic partnership between the Medical Research Council and the University of Oxford.

## Author contributions

The project, methodology, and statistical analyses were developed by AJW, with comment from RC. AJW wrote the paper, with critical review and comment from RC.

## Competing interests

The authors declare no competing interests.

## Footnotes

1 Sir David Cox devised the proportional hazards model, and anticipated several points in this paper. In a 1994 interview he remarked [28], “I would normally want to tackle problems parametrically”, because, “various people have shown that the answers are very insensitive to the parametric formulation of the underlying distribution”.

2 *F* (50)*/F* (100) is the probability of disease by age 50, given that you will have the disease by age 100.

## Notes

### Competing Interest Statement

The authors have declared no competing interest.

## References

[1] Nordling, C. O. A new theory on the cancer-inducing mechanism. British Journal of Cancer 7, 68–72 (1953).

[2] Armitage, P. & Doll, R. The age distribution of cancer and a multi-stage theory of carcinogenesis. British Journal of Cancer 8, 1–12 (1954).

[3] Peto, R. Epidemiology, multistage models, and short-term mutagenicity tests 1. International Journal of Epidemiology 45, 621–637 (2016).

[4] Ai-Chalabi, A. et al. Analysis of amyotrophic lateral sclerosis as a multistep process: a population-based modelling study. Lancet Neurology 13, 1108–1113 (2014).

[5] Chio, A. et al. The multistep hypothesis of ALS revisited the role of genetic mutations. Neurology 91, E635–E642 (2018).

[6] Corcia, P. et al. In ALS, a mutation could be worth two steps. Revue Neurologique 174, 669–670 (2018).

[7] Vucic, S. et al. Amyotrophic lateral sclerosis as a multi-step process: an australia population study. Amyotrophic Lateral Sclerosis and Frontotemporal Degeneration 20, 532–537 (2019).

[8] Licher, S. et al. Alzheimer’s disease as a multistage process: an analysis from a population-based cohort study. Aging-Us 11, 1163–1176 (2019).

[9] Vucic, S. et al. ALS is a multistep process in south korean, japanese, and australian patients. Neurology 94, E1657–E1663 (2020).

[10] Garton, F. C., Trabjerg, B. B., Wray, N. R. & Agerbo, E. Cardiovascular disease, psychiatric diagnosis and sex differences in the multistep hypothesis of amyotrophic lateral sclerosis. European Journal of Neurology 28, 421–429 (2021).

[11] Le Heron, C. et al. A multi-step model of parkinson’s disease pathogenesis. Movement Disorders (2021).

[12] Pearce, N. et al. Does death from covid-19 arise from a multi-step process? European Journal of Epidemiology 36, 1–9 (2021).

[13] Goodnow, C. C. Multistep pathogenesis of autoimmune disease. Cell 130, 25–35 (2007).

[14] Jaiswal, S. et al. Age-related clonal hematopoiesis associated with adverse outcomes. New England Journal of Medicine 371, 2488–2498 (2014).

[15] Jaiswal, S. et al. Clonal hematopoiesis and risk of atherosclerotic cardiovascular disease. New England Journal of Medicine 377, 111–121 (2017).

[16] Mustjoki, S. & Young, N. S. Somatic mutations in “benign” disease. New England Journal of Medicine 384, 2039–2052 (2021).

[17] Alriyami, M. & Polychronakos, C. Somatic mutations and autoimmunity. Cells 10 (2021).

[18] Olafsson, S. & Anderson, C. A. Somatic mutations provide important and unique insights into the biology of complex diseases. Trends in Genetics 37, 872–881 (2021).

[19] Martincorena, I. et al. High burden and pervasive positive selection of somatic mutations in normal human skin. Science 348, 880–886 (2015).

[20] Martincorena, I. et al. Somatic mutant clones colonize the human esophagus with age. Science 362, 911–+ (2018).

[21] Martincorena, I. Somatic mutation and clonal expansions in human tissues. Genome Medicine 11 (2019).

[22] Li, R. et al. A body map of somatic mutagenesis in morphologically normal human tissues. Nature 597, 398–403 (2021).

[23] Burnet, F. M. Auto-immunity and Auto-immune Disease (Medical Technical Publishing Co Ltd (republished 1st edition by Springer), 1972).

[24] Franceschi, C., Garagnani, P., Parini, P., Giuliani, C. & Santoro, A. Inflammaging: a new immune-metabolic viewpoint for age-related diseases. Nature Reviews Endocrinology 14, 576–590 (2018).

[25] Franceschi, C. et al. Inflamm-aging - An evolutionary perspective on immunosenescence, vol. 908 of Molecular and Cellular Gerontology (2000).

[26] Webster, A. J. Multi-stage models for the failure of complex systems, cascading disasters, and the onset of disease. Plos One 14 (2019).

[27] Armitage, P. Multistage models of carcinogenesis. Environmental Health Perspectives 63, 195–201 (1985).

[28] Reid, N. & Cox, D. A conversation with cox,david. Statistical Science 9, 439–455 (1994). URL ://WOS:A1994PX33200010. Reid, n cox, d Reid, Nancy/B-8234-2013.

[29] Nyberg, S. T. et al. Obesity and loss of disease-free years owing to major non-communicable diseases: a multicohort study. The Lancet Public Health 3, e490–e497 (2018). URL https://doi.org/10.1016/S2468-2667(18)30139-7.

[30] Yoshida, K. et al. Tobacco smoking and somatic mutations in human bronchial epithelium. Nature 578, 266–272 (2020).

[31] Bycroft, C. et al. The UK Biobank resource with deep phenotyping and genomic data. Nature 562, 203–209 (2018).

[32] Organization, W. H. International statistical classification of diseases and related health problems 10th revision (2016). URL https://icd.who.int/browse10/2016/en.

[33] Organization, W. H. ICD-11 for mortality and morbidity statistics (icd-11 mms) 2018 version (2018). URL https://icd.who.int/browse11/l-m/en.

[34] Webster, A. J., Gaitskell, K., Turnbull, I., Cairns, B. J. & Clarke, R. Characterisation, identification, clustering, and classification of disease. Scientific Reports 11 (2021). Webster, A. J. Gaitskell, K. Turnbull, I. Cairns, B. J. Clarke, R.

[35] R Core Team. R: A Language and Environment for Statistical Computing. R Foundation for Statistical Computing, Vienna, Austria (2021). URL https://www.R-project.org.

[36] Henningsen, A. & Toomet, O. maxlik: A package for maximum likelihood estimation in R. Computational Statistics 26, 443–458 (2011). URL http://dx.doi.org/10.1007/s00180-010-0217-1.

[37] Terry M. Therneau & Patricia M. Grambsch. Modeling Survival Data: Extending the Cox Model Springer, New York, 2000).

[38] Therneau, T. M. A Package for Survival Analysis in R (2021). URL https://CRAN.R-project.org/package=survival. R package version 3.2-13.

[39] Wasserman, L. All of Statistics (Springer, 2005).

[40] Varrichio, C. grr: Alternative Implementations of Base R Functions (2016). URL https://CRAN.R-project.org/package=grr. R package version 0.9.5.

[41] Dowle, M. & Srinivasan, A. data.table: Extension of ‘data.frame’ (2021). URL https://CRAN.R-project.org/package=data.table. R package version 1.14.0.

[42] Oehlschlägel, J. & Silvestri, L. bit64: A S3 Class for Vectors of 64bit Integers (2020). URL https://CRAN.R-project.org/package=bit64. R package version 4.0.5.

[43] Borchers, H. W. pracma: Practical Numerical Math Functions (2021). URL https://CRAN.R-project.org/package=pracma. R package version 2.3.3.

[44] Nakazawa, M. fmsb: Functions for Medical Statistics Book with some Demographic Data (2021). URL https://CRAN.R-project.org/package=fmsb. R package version 0.7.1.

