## Supplemental Material for "Sporadic, late-onset, and multistage diseases"

### A All diseases are "rare"

All diseases are rare, in the sense that without germline genetic abnormalities or exposures to e.g. highly carcinogenic risk factors such as asbestos, then over most of a typical human lifetime $S(t)$ for any disease is never much less than 1. It is the large numbers of potentially fatal diseases that make living to old ages unlikely, not the inevitability of any single disease. To illustrate the point, if there were only 10 diseases that we could die from, and a 50% chance of surviving each individual disease to age 100, the chance of living to 100 would be $(1/2)^{10}$, approximately 1 in 1000. With 100s of potential causes of death [1] even with a low chance of any individual disease, the chance of living to 100 is very small. As a result, over most of a typical human lifetime it is often reasonable to approximate the probability density function for a disease's incidence as $f(t) = h(t)S(t) \simeq h(t)$. Similarly, for many mathematical purposes, the survival function can be approximated as $S(t) = \exp(-H(t)) \simeq 1 - H(t) \simeq 1$.

This article has considered each individual's first hospital admission for a disease in each ICD-10 chapter. This is intended to approximate an individual's age at first admission to hospital for a disease without confounding by prior disease, from which the cumulative probability function $F(t)$ for disease occurring within age $t$ is estimated for each disease with a Weibull and NSM model. Figure 1 shows the estimated cumulative distribution functions $F(t)$ for all the diseases studied, shown as a histogram of $F(t)$ for ages $45-100$ in 5-year intervals. The figures clearly indicate that all diseases are rare, in the sense that even by age 100, most diseases have less than a 10% chance of having occurred. This justifies the approximations of $F(t) \simeq H(t)$, $S(t) \simeq 1 - H(t) \simeq 1$, and $f(t) \simeq h(t)$.

### B Properties of multistage models

#### B.1 Sequential and non-sequential multi-stage processes

In its most general form, a multistage model can describe any process that leads to disease through one or more independent paths, with one or more independent steps or stages on each path before disease can occur [1]. Throughout we use the usual notation of a cumulative probability $F(t)$, survival function $S(t) = 1 - F(t)$, probability density function $f = dF/dt = -dS/dt$, hazard function $h(t) = f(t)/S(t)$, cumulative hazard function $H(t) = \int_0^t h(s)ds$, and these are related through $S(t) = \exp(-H(t))$ or equivalently $H(t) = -\log(S(t))$.

For diseases that require $m$ independent steps that can occur in any order, then the survival function $S(t)$ is easily calculated in terms of the survival function for each step $S_j(t)$, with [1],

$$S(t) = 1 - \Pi_{j=1}^{m} (1 - S_j(t)) \tag{1}$$

If the hazard rate for each step were the same constant rate $1/L$, then $S_i(t) = exp(-t/L)$, and we have,

$$F(t) = 1 - S(t) = \left(1 - e^{-t/L}\right)^m \tag{2}$$

from which we can obtain $f(t)$ by differentiation. Section B.2 explains why it is reasonable to take an approximately constant rate $1/L$ for all stages. We refer to this very simple non-sequential multistage model as the NSM model, and later fit it to several hundred diseases in the UK Biobank dataset.

The original multistage model of Armitage and Doll imagined mutations occurring at constant rates $1/L_i$ for step $i$, with a sequence of mutations needed to trigger disease. If mutations can occur in any order, then provided $t/L_i \ll 1$,

$$S(t) = 1 - \Pi_{i=1}^{m} (1 - S_i(t)) \simeq 1 - \Pi_{i=1}^{m} (t/L_i) \tag{3}$$

[1] The global burden of disease summarises causes of deaths with over 20 categories, such as cancer, each of which contain numerous subcategories of disease. There are also thought to be several thousand rarely observed diseases, that affect 1 in 10 of the population.

giving $f(t) = -dS/dt \simeq (\Pi_{i=1}^{m} 1/L_i) m t^{m-1}$ which is equivalent to the original result of Armitage and Doll [2].

For diseases that occur through $m$ independent steps that must occur in sequence from $j = 1$ to $j = m$, so that step $j$ is only at risk once step $j-1$ has occurred, then the probability density function is given in terms of the Laplace transforms of the probability density functions for each step, with [1],

$$f(t) = \mathcal{L}^{-1}\left\{\Pi_{j=1}^{m}\mathcal{L}\left[f_j(t_j)\right]\right\} \tag{4}$$

where $\mathcal{L}$ and $\mathcal{L}^{-1}$ indicate the Laplace and inverse Laplace transforms. The original Armitage-Doll model is equivalent to approximating $f_j(t) = h_j(t)S_j(t) \simeq h_j(t)$, and assuming that $h_j = (1/L_j)$ is independent of time. A slightly more general argument would take $f_j(t) \propto (t/L_j)^{m_j}$, and both of these lead to $f(t) \simeq (m/L)(t/L)^{(m-1)}$ for some $m$ and $L$, with the approximation intended to be reasonable over a typical human lifetime. Provided that $t/L \ll 1$, then this is approximately equal to a Weibull distribution that has

$$f(t) = -\frac{d}{dt}\{\exp(-(t/L)^m)\} \tag{5}$$

Here the power $m$ from the model fit is referred to as the "effective" number of steps, and is the number of steps in the model if each step occurred at a constant time-independent rate. Note however that several authors have observed that it will often be reasonable to approximate $h(t)$ as a power of time, which will give a Weibull distribution for values of $t$ beyond the approximation where $f \simeq h$. Therefore we caution that good fits to the incidence data are arguably unsurprising, and need not reflect an underlying multistage process. Several authors have noted that it will often be reasonable to approximate $h(t)$ as a power of time, which will give a Weibull distribution for values of $t$ beyond the approximation where $f \simeq h$. Therefore Weibull models would be expected to fit a much broader range of data than just those generated by multi-stage processes.

#### B.2 Exposures can change the observed number of rate-limiting steps

In the non-sequential multi-stage model of disease, if the rates of one or more of the steps becomes substantially faster than those of the others, then the step will not noticeably influence the incidence data. Mathematically this is clear from Eq. 1, with $(1 - S_j) = 1 - e^{-\mu_j t} \to 1$ for $\mu_j t \gg 1$, as would occur for times with $\mu_{k \neq j} t \sim 1$ and $\mu_j \gg \mu_{k \neq j}$. Therefore, any exposures that substantially increase the rates of one or steps in an NSM disease model, will reduce the number of rate-limiting steps that are observed. An interesting consequence of this argument is that within this non-sequential model, all the rates must be similar, otherwise we will only observe those that are substantially slower. This helps justify the approximation of equal rates for all steps in the NSM model described above.

Now consider diseases that arise through an ordered sequence of stages or steps, as opposed to several steps or stages that can occur in any order. In principle, if an exposure increases the rate of any individual step in the disease process, then the number of steps needed to cause disease does not change. Therefore, in a mathematical model with a sequence of steps needed to cause disease, an exposure might be expected to change parameters that modify disease rate, but not those corresponding to the number of steps $k$. Counter-intuitively perhaps, this is not true. To see why, consider the simplest case with constant hazard rates $\mu_0..\mu_m$ at each step (the Armitage-Doll model), and then we will let $\mu_0$ become large. The probability density function is [1],

$$f(t) = \mathcal{L}^{-1}\left\{\Pi_{j=0}^{m}\mathcal{L}\left[f_j(t_j)\right]\right\} \tag{6}$$

where $\mathcal{L}$ and $\mathcal{L}^{-1}$ indicate the Laplace and inverse Laplace transforms. For the slower rate-limiting steps, then as discussed above, we can approximate $f_j(t_j) \simeq h_j(t_j) = \mu_j$, but for the step whose rate $\mu_0$ will be allowed to increase, we must keep the exact distribution $f_0(t_0) = \mu_0 e^{-\mu_0 t_0}$. Then using standard Laplace transform results with $\mathcal{L}[\mu_j](s) = \mu_j/s$ and $\mathcal{L}[\mu_0 e^{-\mu_0 t}](s) = \mu_0/(s+\mu_0)$,

$$\begin{aligned} f(t) &= \left(\Pi_{j=1}^{m}\mu_j\right)\mathcal{L}^{-1}\left\{\tfrac{1}{s^m}\tfrac{\mu_0}{s+\mu_0}\right\} \\ &= \left(\Pi_{j=1}^{m}\mu_j\right)\mathcal{L}^{-1}\left\{\tfrac{1}{s^{m-1}}\left[\tfrac{1}{s} - \tfrac{1}{s+\mu_0}\right]\right\} \\ &= \left(\Pi_{j=1}^{m}\mu_j\right)\mathcal{L}^{-1}\left\{\tfrac{1}{s^{m-2}}\left[\tfrac{1}{s} - \tfrac{1}{\mu_0}\left[\tfrac{1}{s} - \tfrac{1}{s+\mu_0}\right]\right]\right\} \end{aligned} \tag{7}$$

Taking the inverse Laplace transform either before or after taking $\mu_0 \to \infty$, shows that if $\mu_0$ becomes large then,

$$f(t) \to \left(\Pi_{j=1}^{m}\mu_j\right)\frac{t^{m-1}}{\Gamma(m)}\left[1 + O\left(\frac{t}{\mu_0}\right)\right] \tag{8}$$

as if it were approximated by a model with one less stage. The argument generalises to several steps whose rates become large, or to situations where the rate-limiting steps have a more complex dependence such as $f_j(t_j) \simeq \mu_j t^{m_j}$. This is an unusual example of where the mathematical result is clearer than intuition. It indicates that even if there are several sequential steps in a disease process, they need not be observable in the rate of disease incidence.

Whereas a change in the rate $\mu_j$ or $1/L_j$ can be captured by a proportional hazards model, a change to the effective number of steps $m$ is equivalent to changing the baseline hazard. Therefore anything that changes the effective number of steps for a disease, in principle cannot be studied with a proportional hazards model; although in practice that approximation may often be adequate.

#### B.3 Multistage processes and the proportional hazards assumption

Multistage models highlight some implicit assumptions that are made when we use a proportional hazards model. Firstly consider the simplest multistage process of a non-sequential model with two steps that can occur in any order. In the rare-disease approximation that Assume that for each step that we can approximate $(1 - S_i(t)) = F_i(t) \simeq H_i(t)$, then also using [1] $S(t) = S_1(t)S_2(t)$,

$$\begin{aligned} \log S(t) &= \log\left\{1 - (1 - S_1(t))\,(1 - S_2(t))\right\} \\ &\simeq \log\left\{1 - H_1(t)H_2(t)\right\} \\ &\simeq -H_1(t)H_2(t) \end{aligned} \tag{9}$$

and the hazard $h(t) = -d\log S(t)/dt \simeq h_1(t)H_2(t) + h_2(t)H_1(t)$. Assuming that for each step $h_i(t) = e^{\beta_i X} h_{i0}(t)$, then,

$$h(t) \simeq e^{(\beta_1+\beta_2)X}\left(h_1(t)H_2(t) + h_2(t)H_1(t)\right) \tag{10}$$

which has the proportional hazards form. However, the proportional hazards model will estimate $(\beta_1 + \beta_2)$, that could be very different to $\beta_1$ or $\beta_2$ of steps 1 and 2.

The same is also true for sequential disease processes when each step is sufficiently infrequent that we can approximate $f_i(t) \simeq h_i(t)$ (over the ages observed), and for each step assume that we can write $h_i(t) = e^{\beta_i X} h_{i0}(t)$. For a sequential disease process with two steps, Eq. 16 of Ref. [1] gives,

$$\begin{aligned} f(t) &= \mathcal{L}^{-1}\left\{\mathcal{L}\left[f_1(t)\right]\mathcal{L}\left[f_2(t)\right]\right\} \\ &\simeq e^{(\beta_1+\beta_2)X}\mathcal{L}^{-1}\left\{\mathcal{L}\left[h_{10}(t)\right]\mathcal{L}\left[h_{20}(t)\right]\right\} \end{aligned} \tag{11}$$

that has a proportional hazards form, but as with the non-sequential model, we measure $(\beta_1 + \beta_2)$ and we would not discern differences between $\beta_1$ and $\beta_2$. The important point being made is that multi-stage disease processes could have different risk factors for different stages of the disease, they could even have opposite sign, and we would be unable to discern this from incidence data of a proportional hazards model. The arguments above generalise easily to an arbitrary number of stages.

A related example is a "composite endpoint", a collection of diseases that are studied together because they are thought to have the same underlying biological cause. A composite endpoint of several diseases could have one or more shared steps, and one or more steps that differ. In that example, the proportional hazards assumption will only hold if all diseases have the same risk factors for any steps to disease that differ for diseases in the composite endpoint, or if any different disease processes are sufficiently rapid that they do not change the incidence rate.

#### B.4 The distribution of parameters

The diseases are sufficiently rare that over most human lifespans, the probability density function for disease incidence $f$, can for the purpose of rough estimates be approximated in terms of the hazard function $h$, as $f \simeq h$. As a consequence, the expected number of cases that is proportional to $F(t) = \int_0^t f(s)ds$, has,

$$F(t) \simeq \int_0^t h(s)ds = H(t) \tag{12}$$

where $H(t)$ is the cumulative hazard function, with $H(t) = (t/L)^m$ for the Weibull distribution. For the diseases in our study, there are several 100 to several 1000 cases, differing by about 30% on a log scale. As a consequence, $\log(H) \sim \alpha$, with $\alpha$ equal for all diseases, and as a rough estimate the majority of diseases considered will have,

$$-\log(L) \sim \frac{\alpha}{m} \tag{13}$$

For a Weibull distribution the mean and standard deviation are proportional to $L$. If $m$ is large then disease risk increases rapidly with age, but $L$ will be smaller, making these diseases rarer at younger ages. In contrast, if $m$ is smaller, then $L$ is larger, and the greater standard deviation would be expected to result in these diseases being observed sporadically at ages that are typically younger than those with high $m$. Because we can have $(1/L)(t/L)^{m-1}e^{-t/L} \simeq (1/L)(t/L)^{m-1}$, the observations are also relevant to the non-sequential (NSM) multistage model when $t/L \ll 1$.

### C Competing risks

So-called "competing risks", are most familiar in the context of mortality data, where the observation of one cause of death prohibits the observation of any other. To minimise the risk of confounding by prior disease, this study only included data for the first disease that occurred in each ICD-10 chapter for each individual, and excluded data for any subsequent disease from the same chapter in the same individual. For example, a histogram of data for the age of atherosclerotic heart disease would not include any cases that occurred in individuals after any other hospital diagnosed disease of the circulatory system, such as angina or myocardial infarction. The purpose of this brief summary is to remind the reader why the analysis in this situation of competing risks is correct, and to point out how competing risks require the estimates to be combined if we were comparing results with histograms of observational data.

**Modelling cause-specific data:** In practice it is straightforward to model cause-specific incidence of diseases when there are competing risks. This is clearest if the data are modelled with hazard functions, that describe the probability density for disease in an infinitesimal unit of time, given that the disease has not yet occurred. For example, in terms of hazards $\{h_j\}$, for diseases $j = 1 : m$, the survival function "sub-distribution" [3], for each disease $S_j = \exp\left(-\int_0^t h_j(w)dw\right)$ is what you would observe if there were no competing risks. When disease $j$ prohibits the observation of disease $k \neq j$, the probability density function is $f_j = h_j \Pi_k S_k$, giving a log-likelihood for data $i = 1 : n$, of,

$$\log(L) = \sum_{i=1}^{n} \left( \log\left(h_j(t_i)\right) + \sum_k \log(S_k(t_i)) \right) \tag{14}$$

Derivatives of $\log(L)$ with respect to parameters involving disease $j$ will only involve terms in $h_j$ and $S_j$, so the maximum likelihood estimates (MLEs) are the same as they would be if studied as an individual disease without any competing risks [3].

**Observational data:** Similarly, the hazard function can be used to determine cause-specific survival curves from observational data, such as for Kaplan-Meier estimates. The procedure is valid if the product of hazard rate $h$ and time interval $\delta t$ is sufficiently small that $h\delta t \ll n$, where $n$ is the number at risk at the start of the time interval $t$ to $t + \delta t$. The estimated cause-specific survival curves are the same as the modelled cause-specific survival curves, as was confirmed by the examples shown in this article. However, what you directly *observe*, are data for competing risks, whose histograms and survival functions for any given disease differ from the cause-specific survival curves. Whereas cause-specific survival curves are constructed from estimates of the hazard rate from a specific cause, the observation of death from a particular cause requires survival from the other causes [2]. So if you plotted a histogram for e.g. death from one of several different possible causes, to get the same plot you would need to combine your estimates to account for competing risks, with $f_j(t) = h_j(t)\Pi_k S_k(t)$, and $k$ including all the competing potential causes of death.

### D Supplementary information: Figures

Figure 2 shows an equivalent plot to figure 2 of the main text, that used a Weibull model, but here produced with the NSM model. The plots with both models are very similar. Figure 3 is the same as figure 2 of the main text, but with data for men and women shown separately.

[2] In modern terminology, cause-specific survival curves for mortality are equivalent to "potential", or "counterfactual", outcomes [4, 5].

Figures 4-6 show equivalent plots to figure 4 of the main text that used a Weibull model to fit data stratified by smoking status, but here stratified by diabetes status also, and using an NSM model for comparison. The plots with both models are very similar.

The original Armitage-Doll model imagined disease progressing through a series of stages, leading to the effective number of steps $m$ being an integer. Figures 7 and 8 plot the estimated values of $m$, ordered from smallest to largest. If $m$ were usually integers then we would expect to see a series of steps. There is very little evidence for steps at integer-valued $m$ in the plots of all diseases in men and women (figure 7), but this is unsurprising because only 28 diseases in men and 31 in women had standard deviations of $m$ that were less than $0.25$ (with a confidence interval less than $\simeq 0.5$). There is some appearance of steps near integer-valued $m$ of Weibull fits for non-smokers in figure 8, but the plot only includes diseases that are sufficiently common in smokers for there to be more than $n_{min} = 150$ cases. When the plot was repeated for all diseases in non-smokers with more than $n_{min} = 150$ cases, (as opposed to only diseases that were sufficiently common in both smokers and non-smokers), the steps were no longer visible. Rarer diseases might introduce too much noise into estimates for $m$ to allow any potential steps in $m$ to be visible, or the steps could simply be caused by fewer diseases having a less continuous spread of $m$. Overall, the results suggest that suitable data-stratification is necessary to allow a biologically meaningful determination of $m$.

Smoking might be expected to increase the rates of some disease processes, possibly sufficiently to reduce the effective number of observed steps to disease $m$. There was some evidence for an overall reduction in $m$ in figure 8, but the difference in $m$ between smokers and non-smokers varied substantially between diseases.

### E Supplementary information: Summary data

#### E.1 Number of diseases in study

We considered the 400 most commonly occurring diseases in the UK Biobank dataset of hospital admission affecting men, and 400 that affect women. For more information on the selection of diseases for study, see Ref. [6]. We rejected any diseases for which the numerical fitting algorithm produced invalid covariance matrices (e.g. with infinite entries), or failed to converge within 100 iterations. A $\chi^2$ test was used to compare goodness of fit between Kaplan-Meier estimates for the survival function and the fitted estimates (see main text for more details), taking the variance of the $i$th point from either the Kaplan-Meier estimate $\sigma_0^2(i)$, or from a combination of this and the uncertainty $\sigma_f^2(i)$ in the fitted estimate, giving a total variance $\sigma^2(i) = \sigma_0^2(i) + \sigma_f^2(i)$ for the difference between the two estimates. The former is reported as $p_0$ and the latter as $p_C$, and we only presented results with $p_0 > 0.05$ after an FDR multiple-testing adjustment. Our interest was primarily in multistage processes with $k > 1$, so we only included diseases with $m > 0.8$ that are consistent with $m \geq 0.8$. In the results we showed fits with the NSM model and the adjusted Weibull models, and only included diseases that were present in both sets of data when presenting adjusted results.

The main paper reports results from a Weibull model, a Weibull model that is adjusted for established risk factors, and a non-sequential multi-stage model referred to as the "NSM" model. The number of diseases present at each selection step are shown from left to right in table 1. As mentioned in the main text, 450 of the diseases included with the NSM and Weibull fits were common to both sets, and 335 diseases were common to both the NSM and adjusted Weibull fits. The fit of the adjusted Weibull model involves both covariates and the baseline hazard, and was not compared with the data, or used anywhere in the article. The p-values for those cases are reported as NA (not available), in the table.

Figures 9-10 show example plots of the Weibull model's fit to the cumulative hazard function for men and women, for the diseases with the largest number of cases in the study. These plots are typical.

#### E.2 Table of fitted data - ranked by sporadic versus late-onset

A csv file with the fits for each disease is deposited with the code at the OSF - see link under Data Availability in main text.

**NSM**

| | Iterations <50 | Converged | Adj. $p_C$ >0.05 | Adj. $p_0$ >0.05 | m >0.8 |
|---|---|---|---|---|---|
| M | 85% | 267 | 252 | 246 | 243 |
| F | 86% | 245 | 234 | 228 | 223 |
| Total | 86% | 512 | 486 | 474 | 466 |

**Weibull**

| | Iterations <50 | Converged | Adj. $p_C$ >0.05 | Adj. $p_0$ >0.05 | m >0.8 |
|---|---|---|---|---|---|
| M | 97% | 279 | 268 | 260 | 258 |
| F | 95% | 254 | 248 | 233 | 227 |
| Total | 96% | 533 | 516 | 493 | 485 |

**Adjusted Weibull**

| | Iterations <50 | Converged | Adj. $p_C$ >0.05 | Adj. $p_0$ >0.05 | m >0.8 |
|---|---|---|---|---|---|
| M | 96% | 322 | NA | NA | 200 |
| F | 92% | 325 | NA | NA | 176 |
| Total | 94% | 647 | NA | NA | 376 |

Table 1: For each model considered, the table summarises the number of diseases: for which the numerical model converged and produced covariance matrices with finite values, with no statistically significant differences after an FDR multiple testing adjustment when comparing Kaplan-Meier estimates to the modelled estimates using confidence intervals for both ($p_C > 0.05$), or just the Kaplan-Meier estimate's confidence interval ($p_0 > 0.05$), and with $m > 0.8$.

### References


[1] Webster, A. J. Multi-stage models for the failure of complex systems, cascading disasters, and the onset of disease. *Plos One* **14** (2019).

[2] Armitage, P. Multistage models of carcinogenesis. *Environmental Health Perspectives* **63**, 195–201 (1985).

[3] Collett, D. *Modelling Survival Data in Medical Research* (New York: Chapman and Hall/CRC,, 2014), 3rd edition edn.

[4] Lash, T., VanderWeele, T., Haneuse, S. & K.J., R. *Modern epidemiology* (Wolters Kluwer (4th edition), 2021).

[5] Pearl, J. *Causality* (Cambridge University Press (2nd edition), 2009).

[6] Webster, A. J., Gaitskell, K., Turnbull, I., Cairns, B. J. & Clarke, R. Characterisation, identification, clustering, and classification of disease. *Scientific Reports* **11** (2021). Webster, A. J. Gaitskell, K. Turnbull, I. Cairns, B. J. Clarke, R.

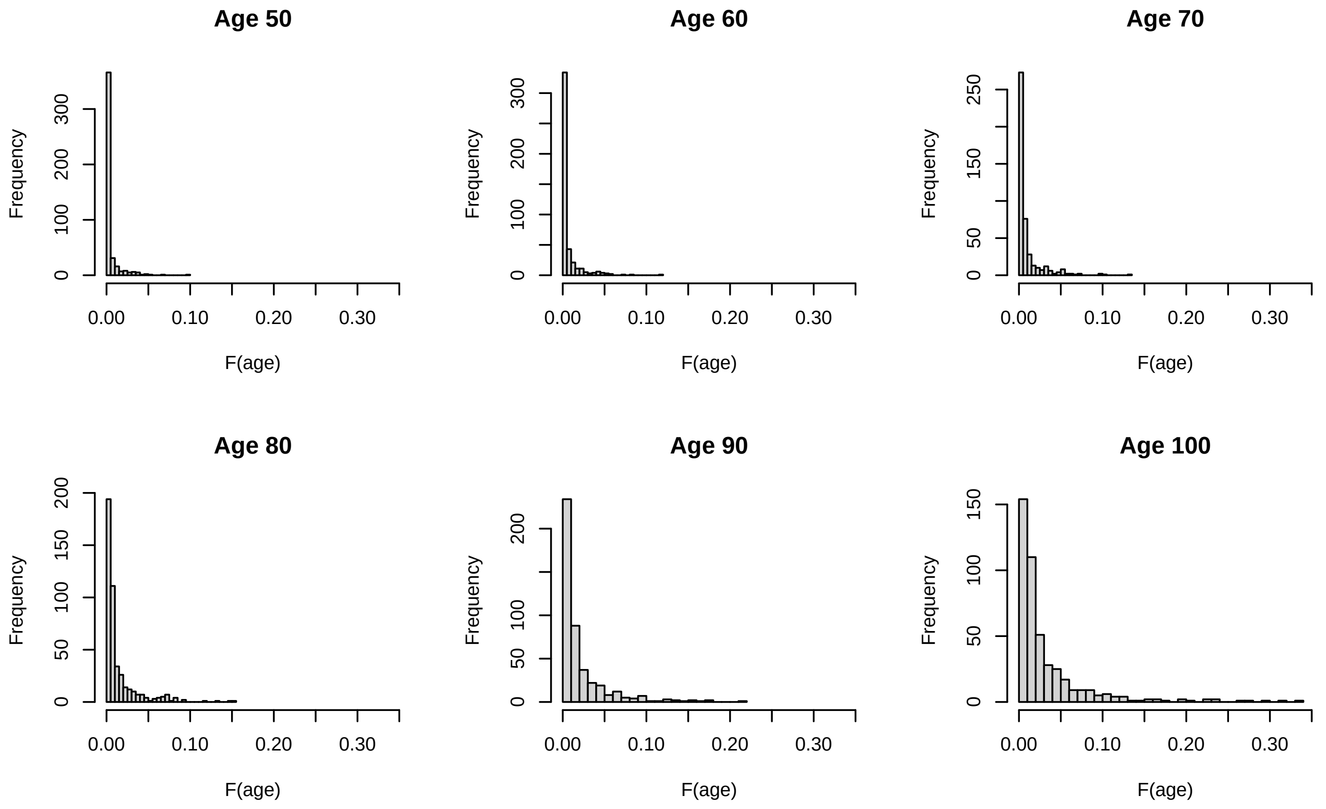


**Weibull model**

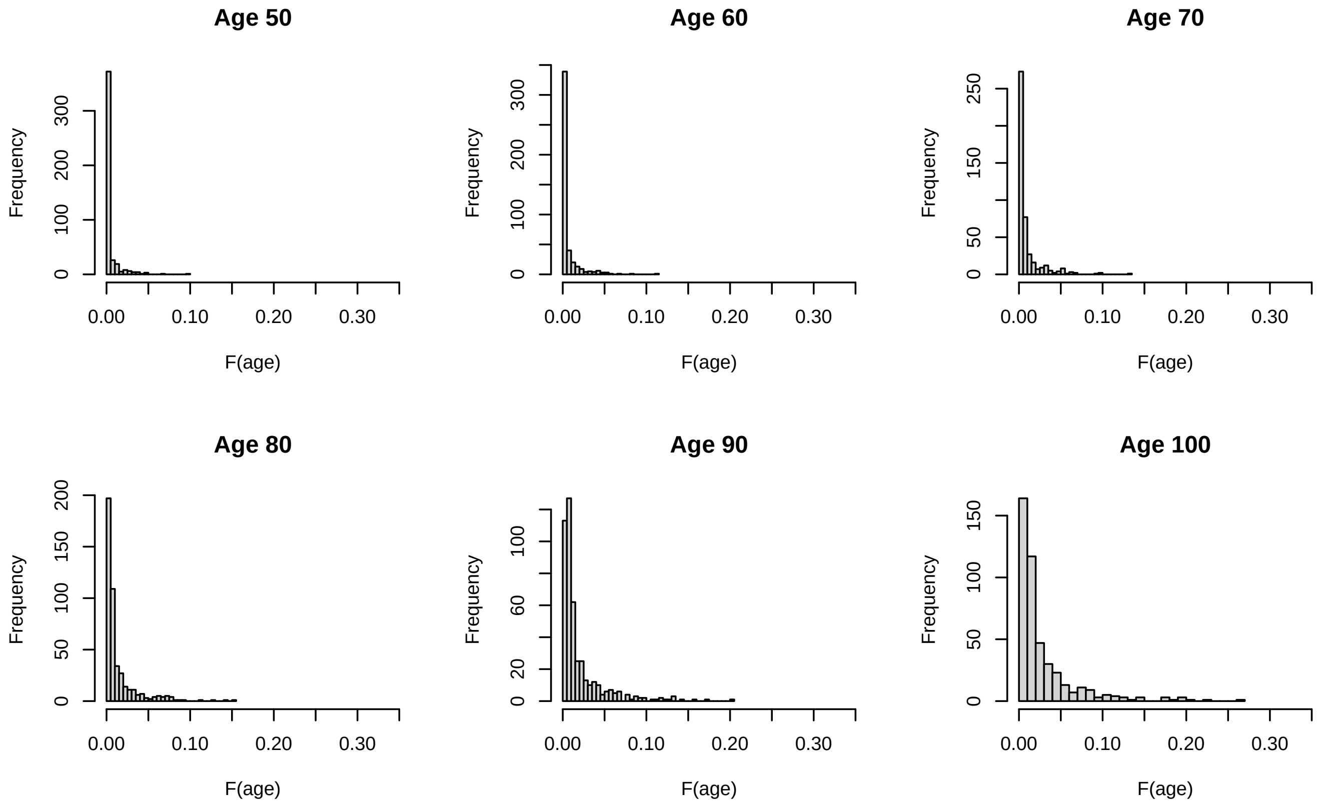


**NSM model**

Figure 1: For each 5-year interval from 45 to 100, and each disease in figure 1 of the main text, a histogram is formed from the estimated c.d.f.s $\{F_i(age)\}$. Even at age 100, the estimated risk of any individual disease (without prior disease) is small.

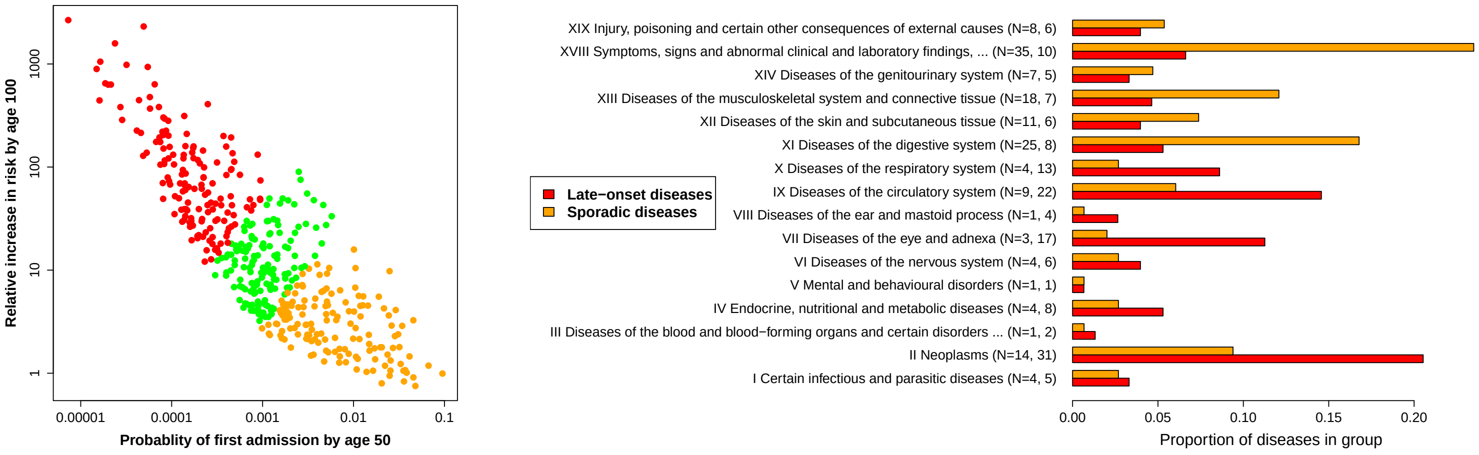


Figure 2: Left: For each disease considered, the probability of first hospital admission is plotted versus the relative increase in probability of disease by age 100. Diseases are split into tertiles to indicate how sporadic, or late-onset, their incidence is (see main text for details). Probabilities were estimated with the NSM model, giving almost identical results to those with the Weibull model in the main text. Right: The composition of sporadic and late-onset diseases is shown in terms of ICD-10 chapters, with N giving the number of diseases.

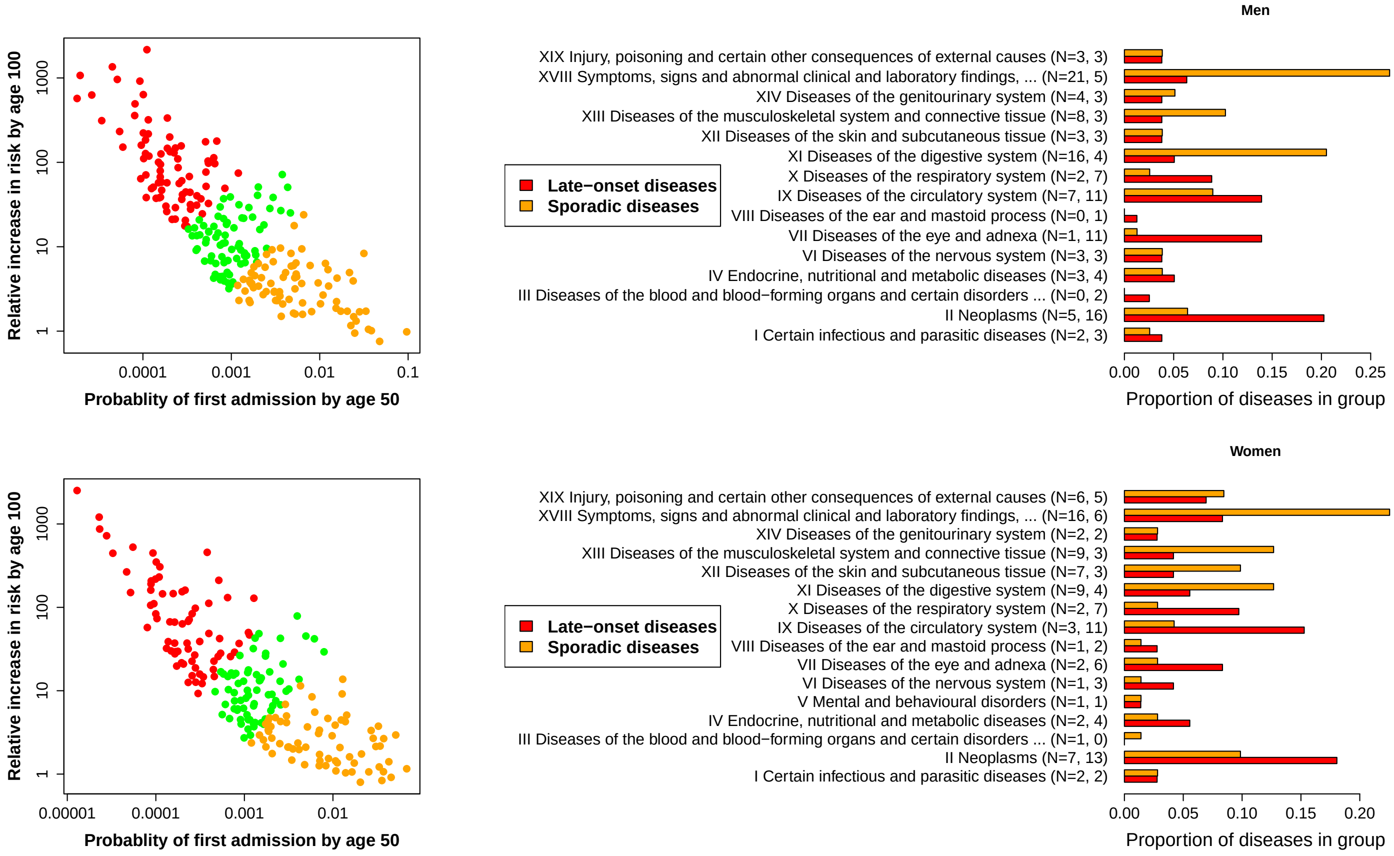


Figure 3: Diseases were considered separately for men (top) and women (bottom). Left: For each disease considered, the probability of first hospital admission is plotted versus the relative increase in probability of disease by age 100. Diseases are split into tertiles to indicate how sporadic, or late-onset, their incidence is (see main text for details). The number and type of diseases studied in men and women are slightly different, leading to slightly different membership of tertiles than when all diseases are considered together. Probabilities were estimated with the Weibull model. Right: The composition of sporadic and late-onset diseases is shown in terms of ICD-10 chapters, with N giving the number of diseases.

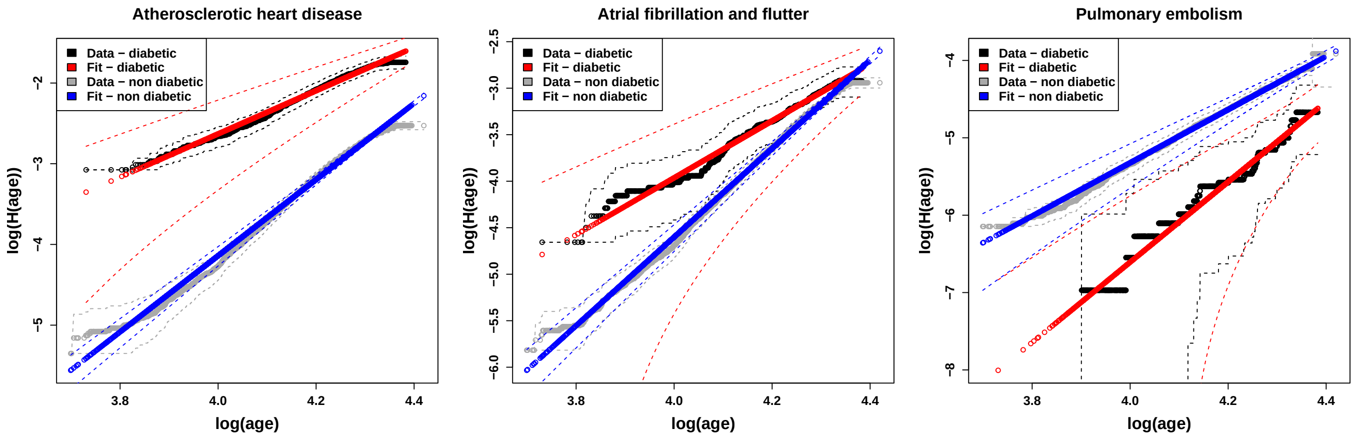


**Men**

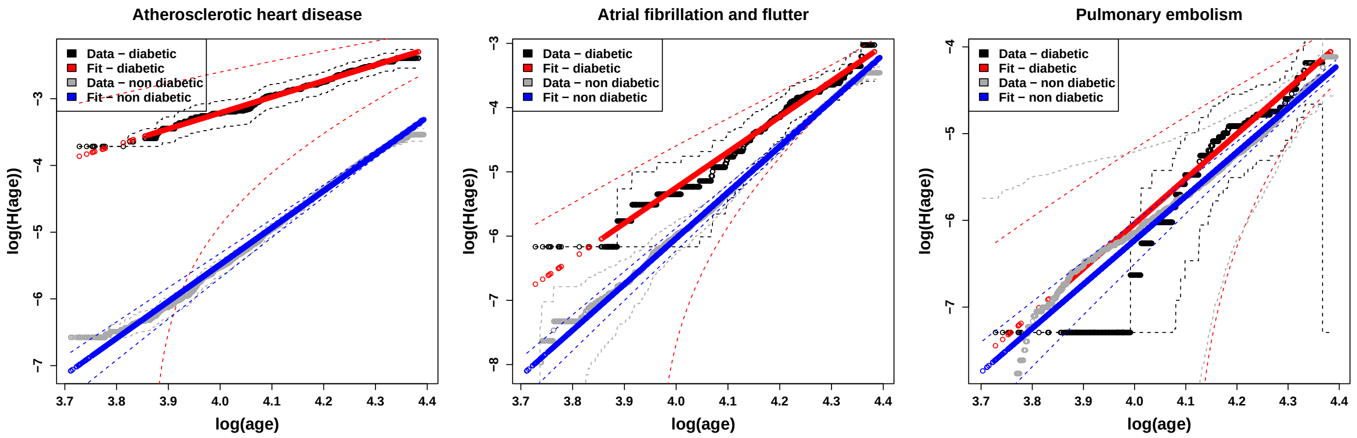


**Women**

Figure 4: The incidence of three common diabetes-related diseases in men (top) and women (bottom), are stratified by diabetes, with Kaplan-Meier estimates adjusted using Eq. 5 of the main text. The data for diabetics differ by vertical displacements and changes in slope, that can be interpreted as increased rates of disease processes that can reduce the numbers of steps needed to trigger disease (see main text for details).

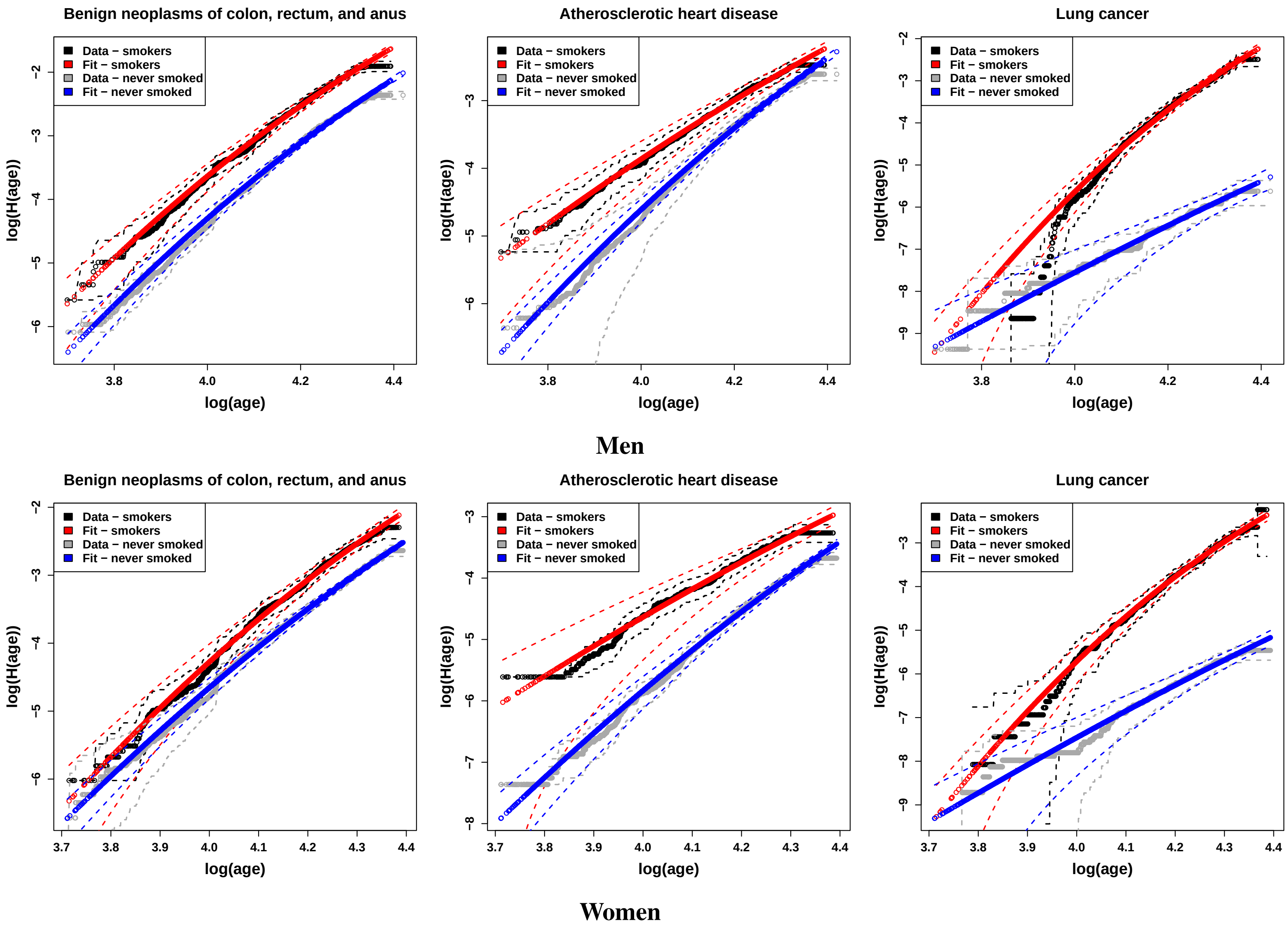


Figure 5: The equivalent to figure 4 of the main text, but calculated using the NSM model. The original caption is repeated here: The incidence of four common smoking-related diseases in men (top) and women (bottom), are stratified by smoking status, with Kaplan-Meier estimates adjusted using Eq. 5 of the main text. The data for smokers differ by vertical displacements and a change in slope. This can be interpreted as increased rates of disease processes, and reduced numbers of steps needed to trigger disease (see main text for details).

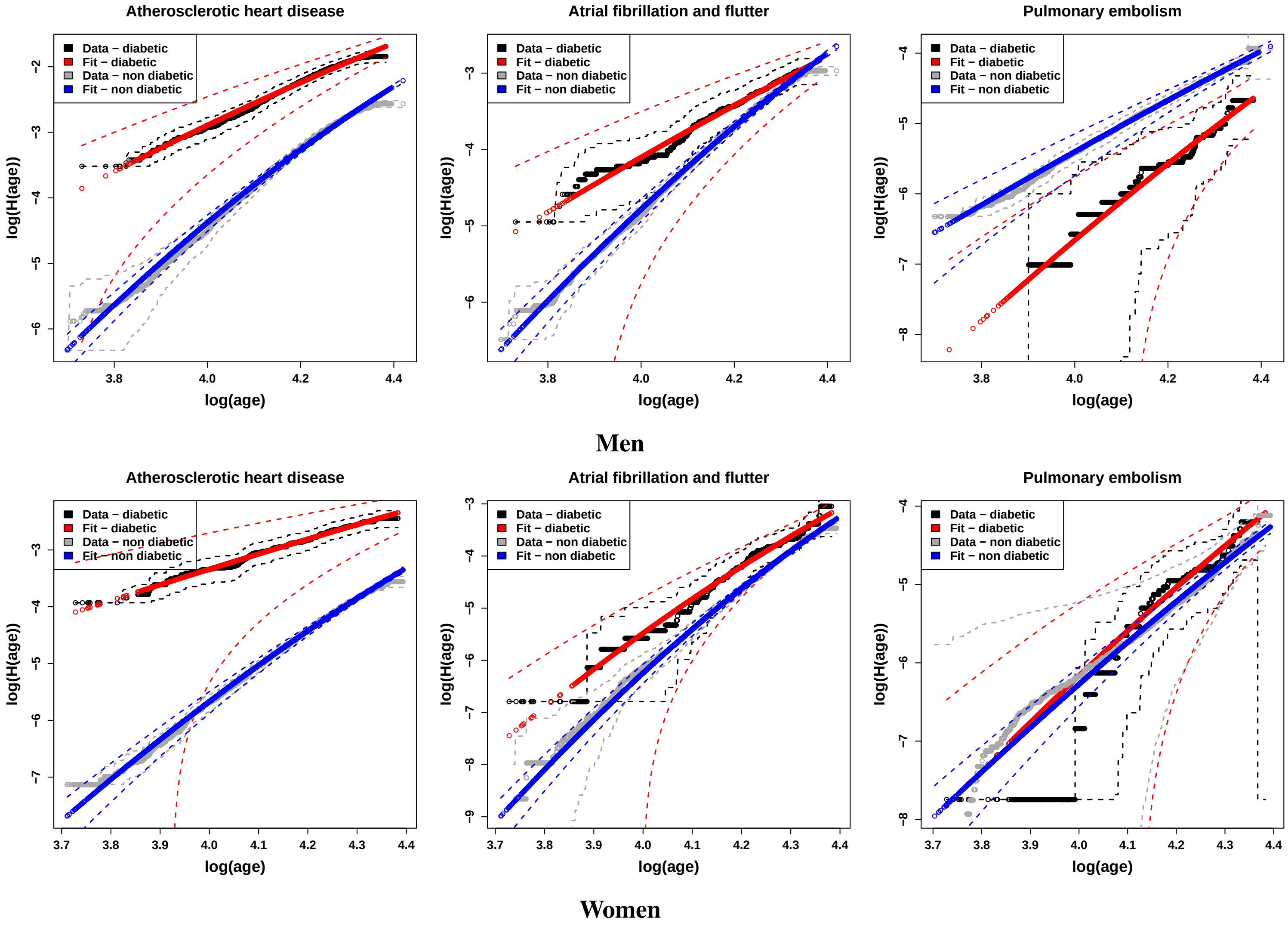


Figure 6: The equivalent to figure 4, but calculated using the NSM model. The original caption is repeated here: The incidence of three common diabetes-related diseases in men (top) and women (bottom), are stratified by diabetes, with Kaplan-Meier estimates adjusted using Eq. 5 of the main text. The data for diabetics differ by vertical displacements and a change in slope. This can be interpreted as increased rates of disease processes, and reduced numbers of steps needed to trigger disease (see main text for details).

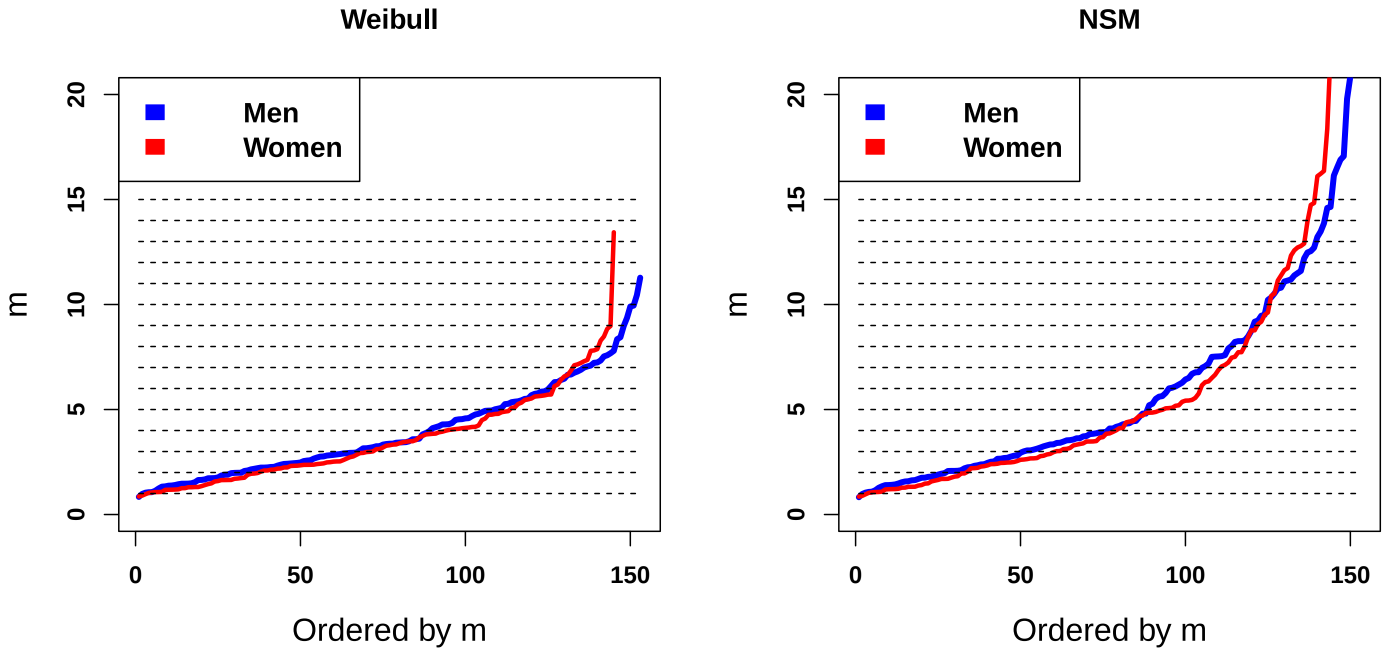


Figure 7: The effective number of steps $m$ needed to trigger disease were estimated for Weibull and NSM models of the same diseases in men and women, and ordered by $m$. There is little visual evidence for steps at integer values of $m$.

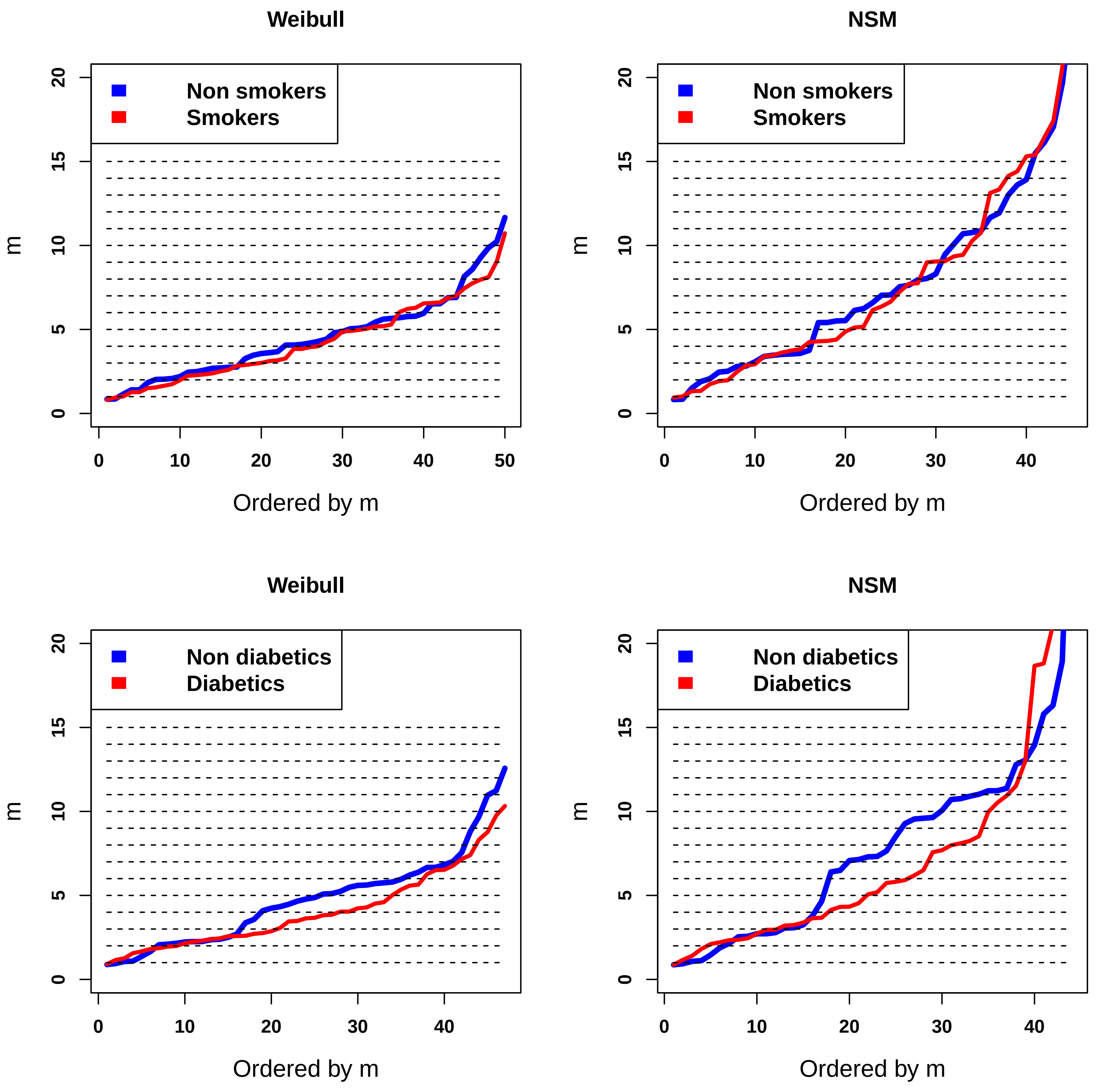


Figure 8: The effective number of steps $m$ needed to trigger disease were estimated for Weibull and NSM models of the same diseases, and ordered by $m$. There is some visual evidence for steps at integer values of $m$ in the Weibull fit for non-smokers. This is discussed further in the supplementary text.

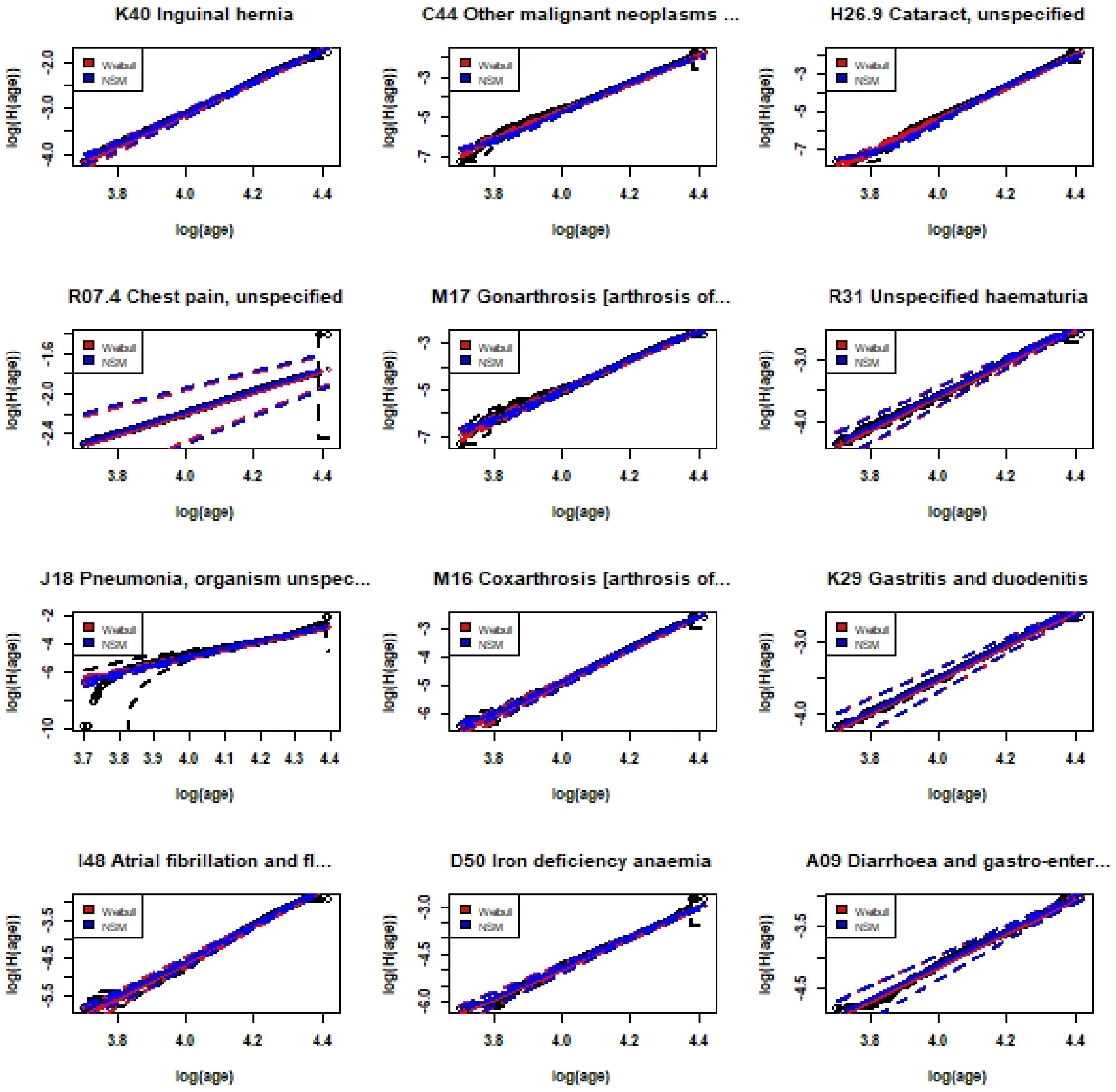


Figure 9: Example plots for the 12 most common diseases in men included in the study. Kaplan-Meier fits are in black, with red (blue) for the Weibull (NSM) models, and 95% confidence intervals estimated by the $\delta$-method indicated by dashed lines. Large confidence intervals can be caused by limitations of the $\delta$-method. Therefore, diseases were selected on the basis of goodness of fit between the modelled estimates and the confidence intervals associated with the Kaplan Meier fits.

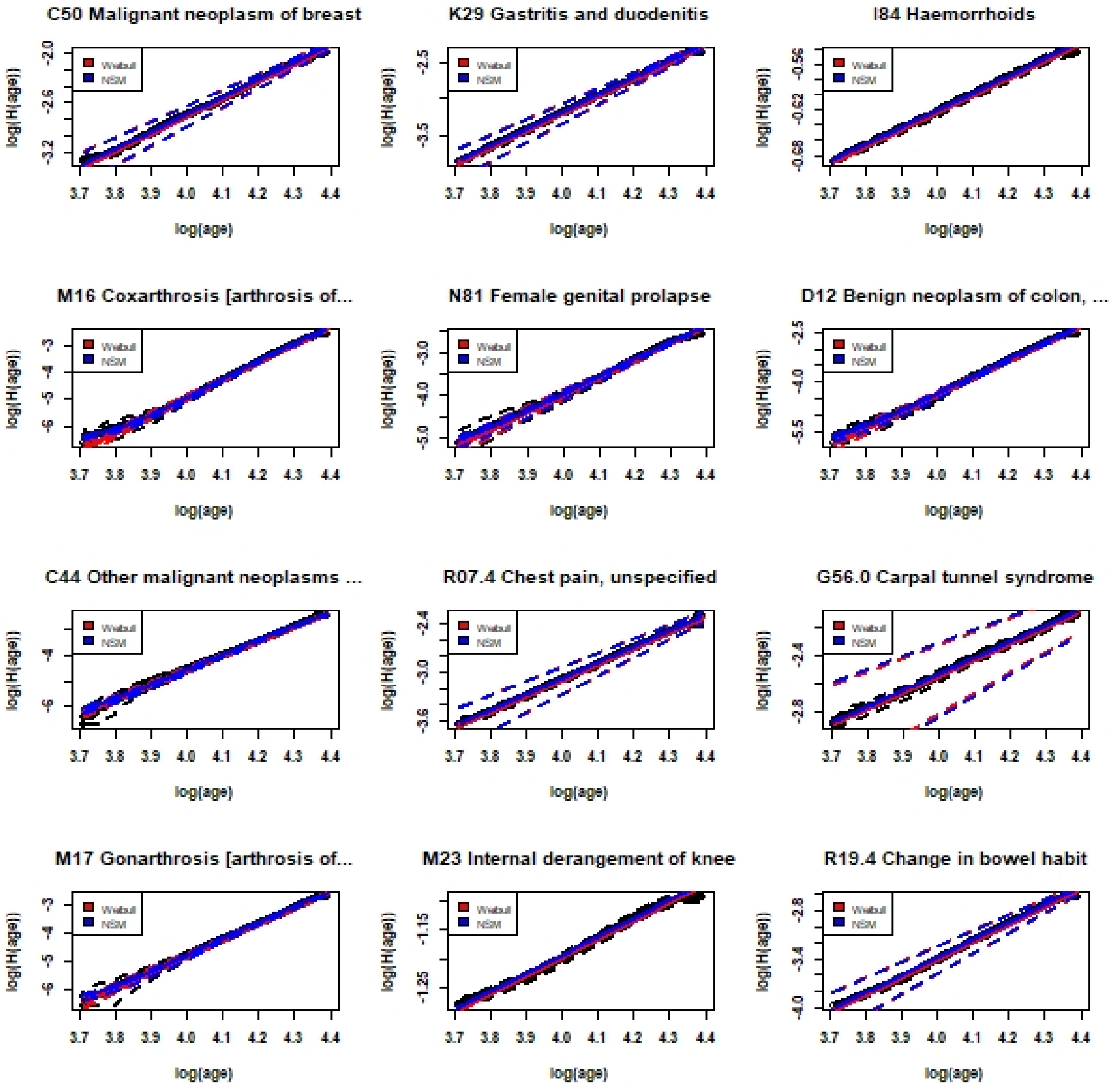


Figure 10: Example plots for the 12 most common diseases in women included in the study. Kaplan-Meier fits are in black, with red (blue) for the Weibull (NSM) models, and 95% confidence intervals estimated by the $\delta$-method indicated by dashed lines. Large confidence intervals can be caused by limitations of the $\delta$-method. Therefore, diseases were selected on the basis of goodness of fit between the modelled estimates and the confidence intervals associated with the Kaplan Meier fits.
